# The Effects of Climate Hazards on Personal Hygiene Practices among People with Disabilities in Bangladesh: A Qualitative Study

**DOI:** 10.1101/2024.12.19.24319192

**Authors:** Shahpara Nawaz, Tasnia Alam Upoma, Arka Goshami, Bithy Podder, Jarin Akter, Mehedi Hasan, Dewan Muhammad Shoaib, Lauren D’Mello-Guyett, Sari Kovats, Mahbub-Ul Alam, Jane Wilbur

## Abstract

People with disabilities experience unique challenges in practising good hygiene, and climate hazards exacerbate those challenges. This study investigates the effects of climate hazards on personal hygiene practices (handwashing, bathing, laundry, and menstrual health) among people with disabilities and their caregivers in cyclone-affected Satkhira and flood-affected Gaibandha of Bangladesh.

A phenomenological qualitative study was conducted in rural Satkhira and Gaibandha. We applied purposive sampling to select 39 people with disabilities and 16 caregivers who experienced cyclones or floods between 2018 and 2023. Data were collected through in-depth interviews, photovoice and ranking, and observations. We thematically analysed findings using Nvivo 14.

During Cyclone Amphan in Satkhira and recurrent floods in Gaibandha, handwashing practices remained unchanged. However, water-logged muddy paths led participants to refrain from bathing for days in Satkhira, causing dissatisfaction about cleanliness. Flooded water points and surroundings in Gaibandha forced participants to bathe and do laundry in contaminated floodwaters, resulting in people reporting increased rashes, skin infections, and fevers. In both districts, the need for bathing and laundering soiled clothes and bedding among people with disabilities experiencing incontinence remained unmet. Additionally, caregivers struggled to provide dignified hygiene support. Women with disabilities could not wash menstrual materials during cyclone in Satkhira. In Gaibandha, contaminated floodwater was used to wash menstrual materials, and individuals living in temporary shelters expressed privacy concerns. Disposal practices were disrupted, with used materials stored indoors or thrown into floodwaters. These challenges adversely affected the health and well-being of people with disabilities and the emotional well-being of the caregivers.

People with disabilities face more complex challenges in maintaining personal hygiene during floods and cyclones due to impairments, gender-specific biological needs, additional health requirements, and dependency on caregivers. To prevent worsening inequalities in a changing climate, it is crucial to systematically include people with disabilities in climate-resilient hygiene initiatives.

## Introduction

Good hygiene practices promote cleanliness, health, dignity, and overall well-being [1]. Hygiene, as defined by the WHO-UNICEF Joint Monitoring Programme (JMP), encompasses conditions and behaviours that help maintain health and prevent disease [2]. Globally, poor hygiene contributes to the burden of infectious diseases, including diarrheal and respiratory infections [3–7], reproductive tract and urinary tract infections [8–11], neglected tropical diseases [12–14], and skin infections [3, 5, 15, 16]. These issues often result in social stigma, emotional distress, and mental health challenges [17, 18]. Inadequate access to Water, Sanitation and Hygiene (WASH) services exacerbates these problems all over the world, particularly in low- and middle-income countries (LMICs) [17, 19–21]

The US Centers for Disease Prevention and Control defines personal hygiene as “regular washing of parts of the body and hair with soap and water (including washing hands and feet), grooming nails, facial cleanliness, covering coughs and sneezes, and menstrual hygiene” [22]. Menstrual health has been defined in more detail as “accessing accurate information about the menstrual cycle, self-care and hygiene behaviours, effective and affordable menstrual materials, water and soap to wash the body and material used with private spaces to do so, disposal mechanisms, medical support for any menstrual related discomfort or disorders and a positive environment free from stigma to enable participation in all aspects of life” [23]. Additionally, managing incontinence - a condition involving the involuntary sporadic or regular loss or leakage of urine or faeces - requires attention to prevent skin infections, smell, and urinary and bladder complications [24].

Access to water for bathing, handwashing, laundry, menstrual health and managing incontinence is vital for hygiene and well-being [25–30]. However, climate variability and change affects personal hygiene practices by threatening access to and continuity of water sources, sanitation and hygiene facilities [31, 32]. Climate change- induced extreme weather events cause infrastructure damage, water contamination [33–37]; drought-induced water shortages [38–41]; salinisation [42, 43]; disruption of supply chains of hygiene items [3, 44], and destruction of sanitation infrastructure [36, 37, 45, 46]. All of which increase the risk of infectious diseases due to declining sanitation and hygiene practices [3, 5, 37, 44, 47–49].

People with disabilities are disproportionately affected by climate hazards, facing barriers to accessing WASH facilities [30, 50], expressing needs [51–53]; receiving critical safety information during climatic events [32, 54]; and insufficient inclusion and accessibility in preparedness and response efforts [55, 56]. Those managing incontinence or menstruating experience heightened challenges, including inadequate WASH facilities, inaccessible information and limited menstrual materials [25–28, 30, 57]. Additionally, caregivers who assist those with disabilities rarely have support or guidance about how to carry out that role hygienically and with dignity [58–60].

Bangladesh, highly vulnerable to climate change, has a total population of over 165 million, with 90 million people in “high climate exposure areas” [37, 61, 62]. With 8% of the country’s total population (approximately 13.2 million) living with disabilities [63], a substantial proportion of them are likely to reside in these climate-exposed areas. Despite significant risks to their health and well-being, research on the effects of climate change on WASH services and how this affects people with disabilities remains limited [64, 65], particularly regarding climate resilience for their hygiene management. Furthermore, national strategies such as the National Adaptation Plan of Bangladesh (2023-2050) [66], the National Strategy for Water Supply and Sanitation (2014) [47], and the Bangladesh Climate Change Strategy and Action Plan 2009 [67] emphasise climate change adaptation and improving WASH service resilience. Similarly, the Persons with Disabilities Rights and Protection Act (2013) focuses on disability rights [68]. However, it is unclear whether these legislations incorporate specific directives for enhancing climate resilience, particularly regarding hygiene, and adequately address the hygiene needs of people with disabilities. A recent analysis of Bangladesh’s WASH policies and guidance highlights that while there is an intent to ensure access for persons with disabilities, documents lack well-defined and resourced activities to achieve this [69].

Our study examines how climate hazards affect the ability of people with disabilities and their caregivers to practice personal hygiene in Satkhira and Gaibanda districts, Bangladesh. Our two research questions are: 1) How do climate hazards affect women and men with disabilities’ personal hygiene practices? 2) How do individuals with disabilities adapt their personal hygiene practices in response to climate hazards, and what are the effects on them and their caregivers?

## Methodology

### Study design

This study employs a phenomenological qualitative research design to explore participants’ thoughts, perceptions and practices regarding maintaining personal hygiene during and immediately after two hydro-meteorological hazards (cyclones and floods).

### Study sites

We selected Satkhira and Gaibandha districts based on Bangladesh’s multi-hazard risk level assessments [70]. Satkhira, located in the southern coastal region, faces cyclones, sea level rise, and saltwater intrusion [54, 71], while Gaibandha, in the northern inland region, experiences frequent flooding and river erosion [70]. These regions highlight diverse climate risks, allowing us to study their effects on the hygiene of people with disabilities.

### Study population and sampling method

Our study included individuals with disabilities (aged 15+) and their caregivers who had experienced a cyclone or flood within the last five years. Participants were purposively selected from rural Satkhira and Gaibandha using World Vision Bangladesh lists documenting members’ names, types of impairments, age, gender, and geographic location. We also used snowball sampling to ensure representation across these variables. People with disabilities were identified using the Washington Group Short Set of questions [72]. Those who reported ’a lot of difficulty’ or more in any functional area were classified as having a disability. Caregivers were selected if they supported the person with disabilities’ personal hygiene. Participants were recruited between 23 August and 31 November 2023

We collected qualitative data with 39 individuals with disabilities and 16 caregivers (Table 1). Among people with disabilities, 17 were female and 22 were male, representing a balanced gender distribution. Participants varied in age, with the majority falling between 18 and 64 years. Participants exhibited a range of impairments, most with multiple impairments (20), followed by mobility impairments (16). All 16 caregivers were female. Most supported individuals with multiple impairments (12), which included people with cognition and self-care; mobility and self-care, and cognition, communication and self-care limitations.

**Table 1.** Study population characteristics.

| Study population | Characteristics | N |
| --- | --- | --- |
| People with disabilities | **Gender** |  |
|  | Female | 17 |
|  | Male | 22 |
|  | **Age group** |  |
|  | 15-17 | 7 |
|  | 18-30 | 16 |
|  | 31-64 | 15 |
|  | 65 and above | 1 |
|  | **Impairment type** |  |
|  | Visual | 3 |
|  | Hearing | - |
|  | Mobility | 16 |
|  | Cognition | - |
|  | Communication | - |
|  | Self-care | - |
|  | Multiple<sup>a</sup> | 20 |
|  | **Location** | - |
|  | Satkhira | 17 |
|  | Gaibandha | 22 |
| **Female Caregivers** | Functional domain of the person with disabilities that the caregiver is supporting |  |
|  | **Impairment type** |  |
|  | Visual | - |
|  | Hearing | - |
|  | Mobility | 4 |
|  | Cognition | - |
|  | Communication | - |
|  | Multiple<sup>a</sup> | 12 |
|  | **Location** | - |
|  | Satkhira | 7 |
|  | Gaibandha | 9 |

<sup>a</sup>Multiple refers to when participants experience limitations across more than one functional domain

### Data collection methods

We applied three qualitative data collection tools to enable methods triangulation: in-depth interviews, observation, and photovoice and ranking (Table 2). We developed separate topic guides (S1-S4) by drawing on the existing evidence at the intersection of climate resilience, disability, and WASH, as well as the research expertise of the team members. Themes explored the personal hygiene practices of people with disabilities and the hygiene support of caregivers during climate hazards. Examples of these include ‘challenges in accessing and using bathing, laundry, and handwashing facilities’, ‘challenges in menstrual health and incontinence management’, ‘management strategies applied by people with disabilities and their caregivers’, and the ‘effects of the challenge on health and well-being’ and ‘effects of the management strategies on health and well-being’. The research team repeatedly reviewed and refined the topic guides, testing them with participants in Satkhira and making further modifications based on the feedback received. We continued to adapt them to explore emerging themes during data collection. The topic guides can be found in supplementary files (S1-S4).

**Table 2.**
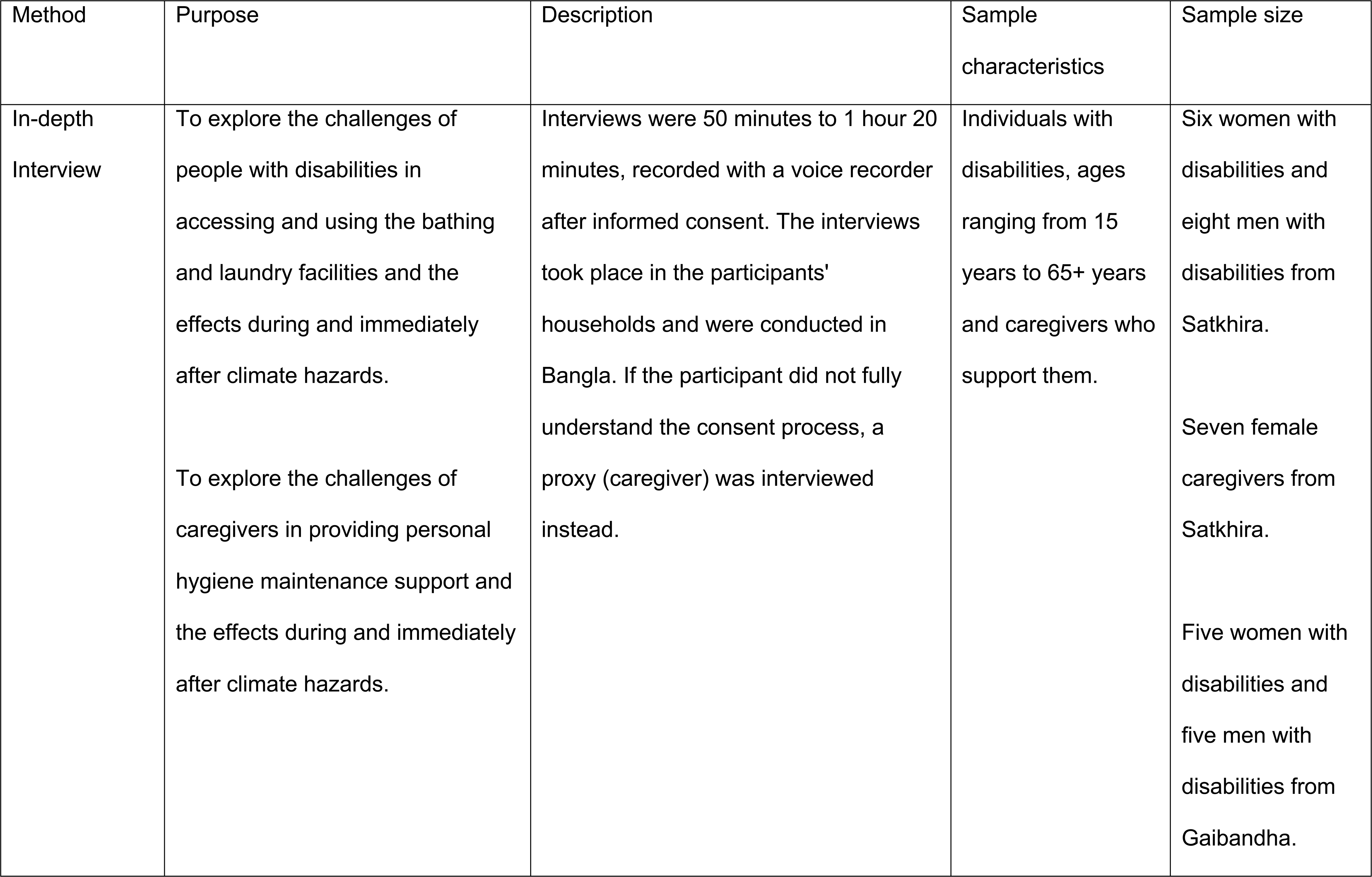
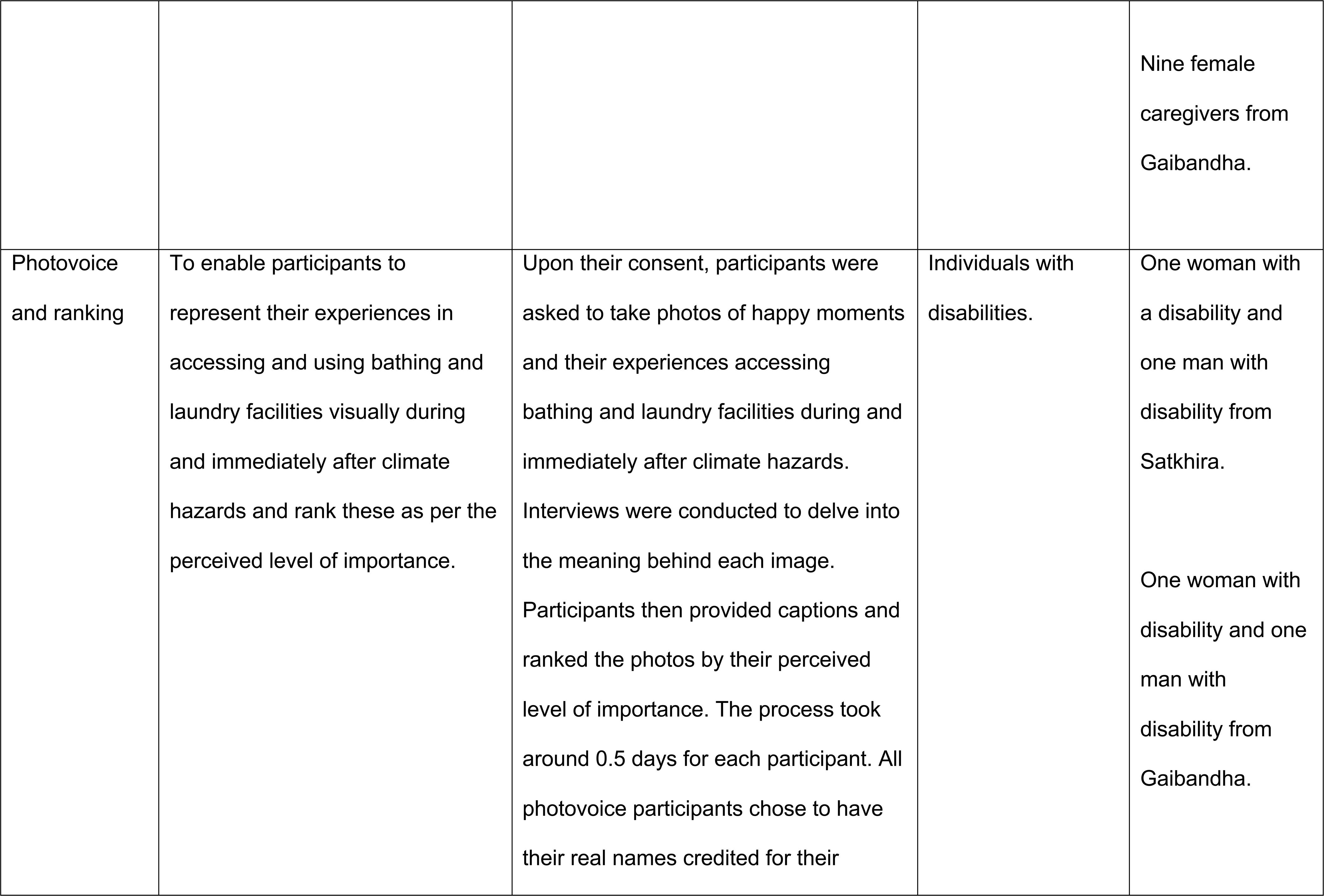
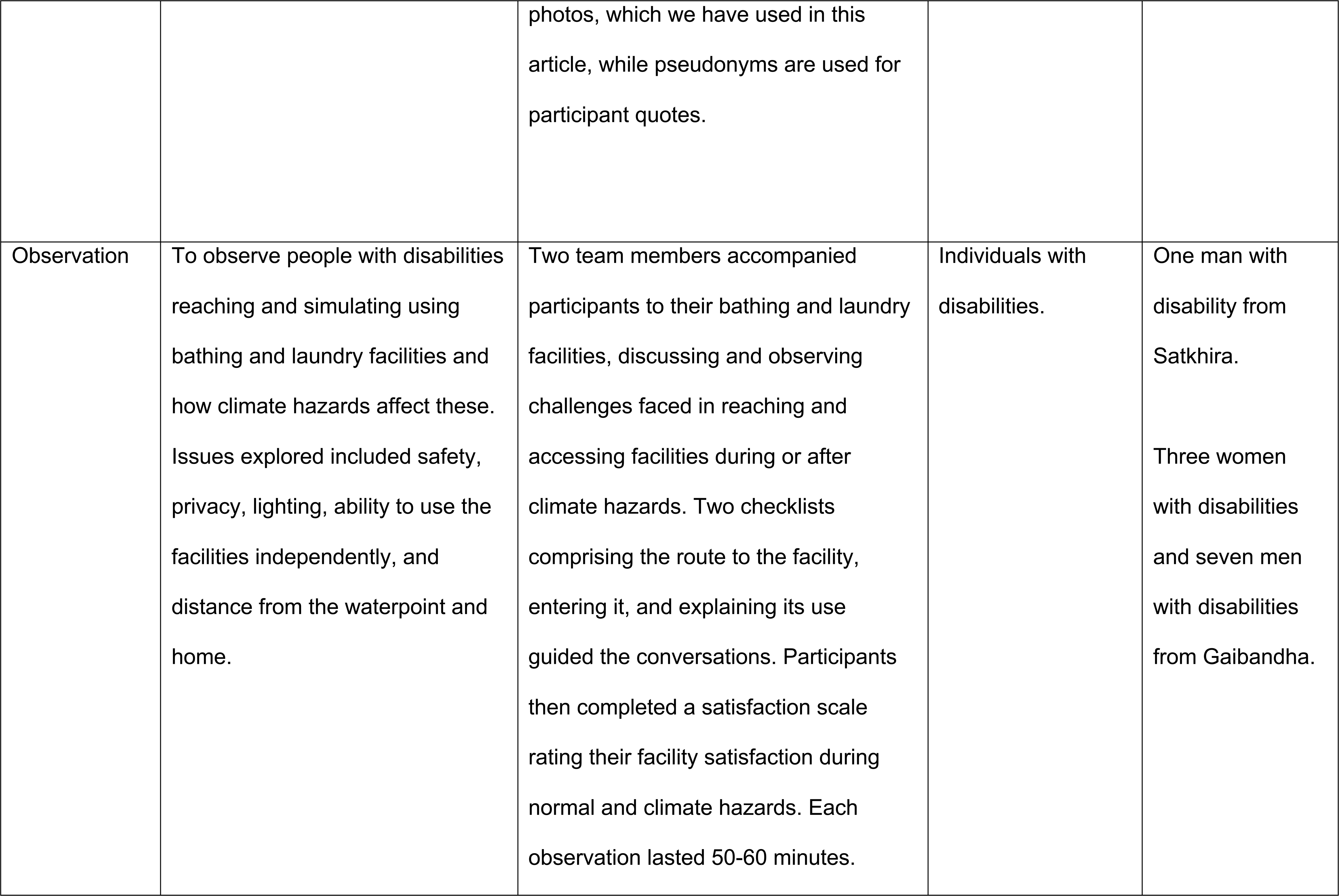
Overview of qualitative data collection methods and sample size.

### Data management and analysis

The International Centre for Diarrhoeal Disease Research, Bangladesh (icddr,b) and the London School of Hygiene and Tropical Medicine (LSHTM) collected data between August and October 2023. The interviews were conducted in Bangla and recorded with the interviewee’s consent. Each day, the researchers reviewed field notes and identified emerging themes. The Bangla-speaking research team members translated and transcribed interview transcripts into English and checked the transcriptions against the original recordings to ensure accuracy and quality.

We used an iterative thematic analysis approach, starting with developing a codebook of a priori codes. Coding initial transcripts and reviewing field notes enabled us to add inductive codes (e.g. ‘availability of water’, ‘managing menstrual product’, and ‘caregiver’s incontinence support’) in the codebook to capture any new insights. Transcripts were coded using NVivo 14 (Lumivero, Colorado, USA), and we assessed inter-coder and intra-coder reliability to ensure consistency and resolve coding disagreements. We created a data display matrix to compare and contrast the coded data on a case-by-case basis, identifying key themes.

We also identified quotes pertinent to each of the themes and captured those in this paper. We summarised the data for each theme, triangulating findings from the three data collection methods. Finally, we reviewed the summaries and compiled the results for each thematic area, which are presented in this paper. To ensure anonymity, quotes were presented excluding names of participants.

### Ethical considerations

We obtained ethical approval for the study from the ethics review board of icddr,b (reference 23072) and the LSHTM (reference 28925). Before starting data collection, we sought and received written informed consent from all participants. For participants under 18 years or those with intellectual disabilities who could not fully understand the consent process, we used simplified information sheets to obtain assent and sought consent from their caregivers.

Caregivers were then interviewed as a proxy, but efforts were made to involve participants directly. Information sheets were read aloud to explain the study’s purpose, procedures, benefits, risks, confidentiality, and the right to refuse or withdraw from the study. We conducted the interviews privately in Bangla, ensuring participants that their information would remain confidential and anonymous in compliance with Bangladeshi law.

Only adults participated in the photovoice and ranking exercises, and they completed two informed consent processes. Initial written informed consent was obtained before participants took photos. During the initial consent process, we explained the purpose of the study and what we would ask people to do if they agreed to participate. A second written informed consent was sought after participants had taken the photos and before we interviewed them about the images. It was explained to the participants that they are the owners of the photos. Participants also identified how they wanted their photos to be used (e.g., in research articles, workshops, or presentations) and if they wanted their real name or pseudonyms credited when used. They could also decide whether their faces should appear blurred or visible. We also sought their consent for any third parties captured in the photos, allowing them to choose if they wanted their faces blurred or shown.

## Results

### Climate hazards experienced

Most participants reported experiencing cyclone and floods (climate hazards) between 2018- 2023. In Satkhira, all referenced hygiene challenges during and after Tropical Cyclone Amphan (2020). Cyclone Amphan struck Satkhira along with eight other southern districts of the country, causing 26 deaths and affecting 2.6 million people [73]. In Gaibandha, participants described challenges in managing hygiene during and after severe floods, with experiences linked to floods in 2019, 2020, 2022 and 2023. Each 2019, 2020, and 2022 flooding event affected over five million people, causing 114, 257, and 141 deaths, respectively, and affecting multiple districts [73]. Across both districts, cyclone and floods were cited as major factors reducing access to WASH facilities and causing disruption in their hygiene practices [37, 74].

### Handwashing

Handwashing practices did not change during or immediately after climate hazards for most participants. Some participants used tube well water for handwashing, occasionally with soap or ash. Even without the additional constraints introduced by the climate hazards, many people with disabilities found independent handwashing difficult, requiring caregiver support. A caregiver explained her usual routine for assisting her daughter:

*“I give water in front of her in a bowl, and I also pour water on her hands, then she washes her hands in the bowl.* (Caregiver, Satkhira)”

During climate hazards, caregivers continued assisting by bringing small bowls of water for handwashing:

*“I used a bowl…I gave her a bowl with water during cyclone… she washed her hands there because she cannot go outside…*” (Female with disability, Satkhira)

Due to affordability issues, most participants could not purchase soap or detergent, so they used ash as an alternative cleaning agent during normal times and climate hazards.

*“Soap is not always available. Sometimes, I wash my hands with ash after returning from the toilet….”* (Male with disability, Satkhira)

### Bathing and laundry

Most participants bathed and did laundry independently or with caregiver assistance at tube wells near their homes under normal conditions. However, accessing these tube wells became challenging during or after Cyclone Amphan in Satkhira and prolonged floods in Gaibandha, due to waterlogged paths and submerged tube wells. In rural areas, we observed that the soil paths to tube wells were often narrow and uneven. During climate hazards, they became muddy, slippery, or water-logged:

*“There was water on the road during [cyclone] Amphan… There was so much mud in the road towards the tube well that I was in the mud up to my knees.”* (Caregiver, Satkhira)

Some participants refrained from bathing during periods of cyclones and flooding and expressed dissatisfaction with their inability to bathe, as described by Liton: *“During rainy days [during the cyclone] I waited for the area to dry…I did not take a bath in these 3 to 4 days. When I cannot shower for 3-4 days, I feel helpless and bad.” (Male with disability, Gaibandha)*

Participants also struggled with the inadequate water available for bathing. In Satkhira, some relied on limited rainwater. A caregiver from Satkhira whose daughter has multiple impairments (Fig 1), described the lack of access to the water point during the cyclone and that she lacked stored water at home. She resorted to collecting rainwater in a bucket and used that small amount to intermittently bathe her daughter.

**Fig 1.**
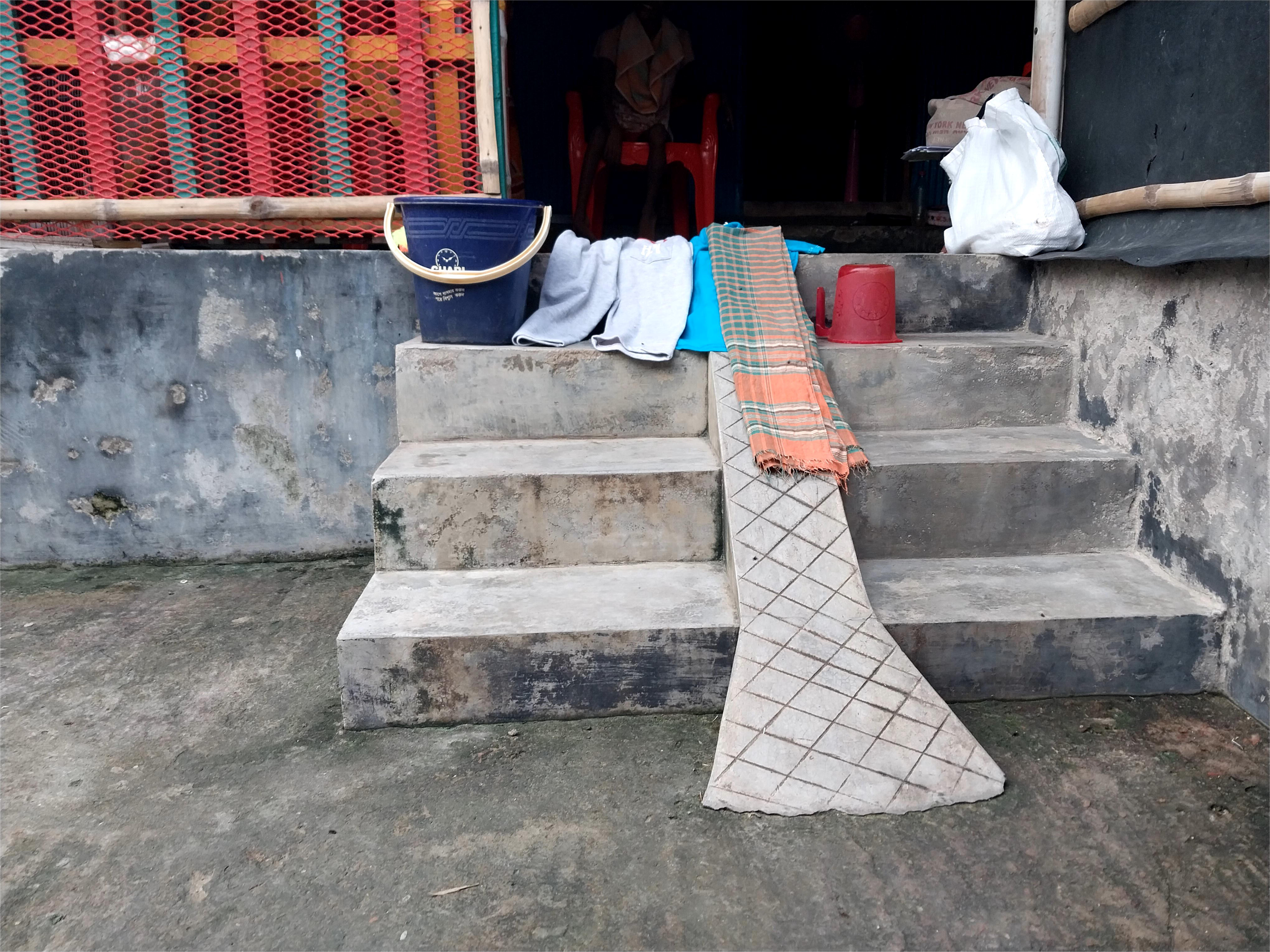
“I had to bathe my daughter during the cyclone Amphan with the little water I collected from the rainfall because I did not have water in my house.” (Caregiver, Satkhira).

During the floods in Gaibandha, tubewells and access routes were submerged in floodwater that was likely contaminated with urine and faeces due to overflowing latrines and widespread open defecation. Unable to access clean water, many participants resorted to bathing in the contaminated floodwaters surrounding their households.

*“There was water everywhere… at that time I used flood water for our bathing…I was unable to reach the tube well traversing through the flood water…so I took a bath in the flood water.”* (Female with disability, Gaibandha)

Furthermore, the flooded surroundings in Gaibandha did not allow participants to reach water points to collect clean water for laundry. As a result, they used contaminated floodwater.

*“I washed them (laundry) in flood water. The portion of the flood water that seemed clean, I washed clothes there. I didn’t have any other option. Where could I wash the clothes when there is water everywhere?”* (Caregiver, Gaibandha)

Participants from Gaibandha reported health issues related to bathing and doing laundry in flood water: *“During the flood, we faced rash and itching on our skin…I just applied a mixture of mustard oil and turmeric to my affected area.”* (Male with disability, Gaibandha)

A male with disability who experienced flood, explained that he believed frequent bathing in floodwaters affected his health, resulting in a fever lasting several days. *“I took a bath on the road with flood water during the flood of August 2018… I suffered from fever for three days during that flood because of bathing in flood water.”* (Male with disability, Gaibandha)

### Managing incontinence

The challenges with bathing and laundry were heightened for people with disabilities experiencing incontinence. Most lacked incontinence products (e.g. absorbent underwear or mattress protectors), resulting in frequent soiling of clothes and an increased need for regular bathing and laundry, even during normal circumstances. During and after cyclones and floods, the inability to access toilets led to increased defecation and urination on clothes and bedding, further heightening the demand for frequent bathing. However, this need often went unmet, compromising cleanliness and well-being. In Satkhira, many people could not bathe as needed and relied on caregivers, who faced challenges maintaining their privacy and dignity.

“.*..he defecated in the room that night… After the storm had stopped the next morning, I got water from the tube well with a bucket and threw the water on him, keeping him seated on the balcony.”* (Caregiver, Satkhira)

In Gaibandha, doing laundry for those with incontinence was even more difficult during the floods. Clothes and bedding soiled with urine and faeces posed a health risk, but frequent washing was significantly challenging. Caregivers had to wash soiled clothes in floodwater, which increased the risk of exposure to water-borne diseases. Washing soiled clothes and bedding in contaminated floodwater posed significant health risks for both caregivers and individuals with incontinence.

### Menstrual health

Eight women with disabilities in Satkhira and Gaibandha reported menstruating during and after cyclones and floods. Most of them used old, worn-out pieces of folded cloth due to affordability issues with disposable or reusable menstrual products. Typically, menstrual cloths were washed with soap, detergent, and tube well water, but cyclones and floods disrupted access to tube wells, stored water at home and outdoor drying. For instance, A caregiver from Satkhira (Fig 2) had to store her daughter’s used menstrual materials indoors and wash or dispose of them after the storms.

**Fig 2.**
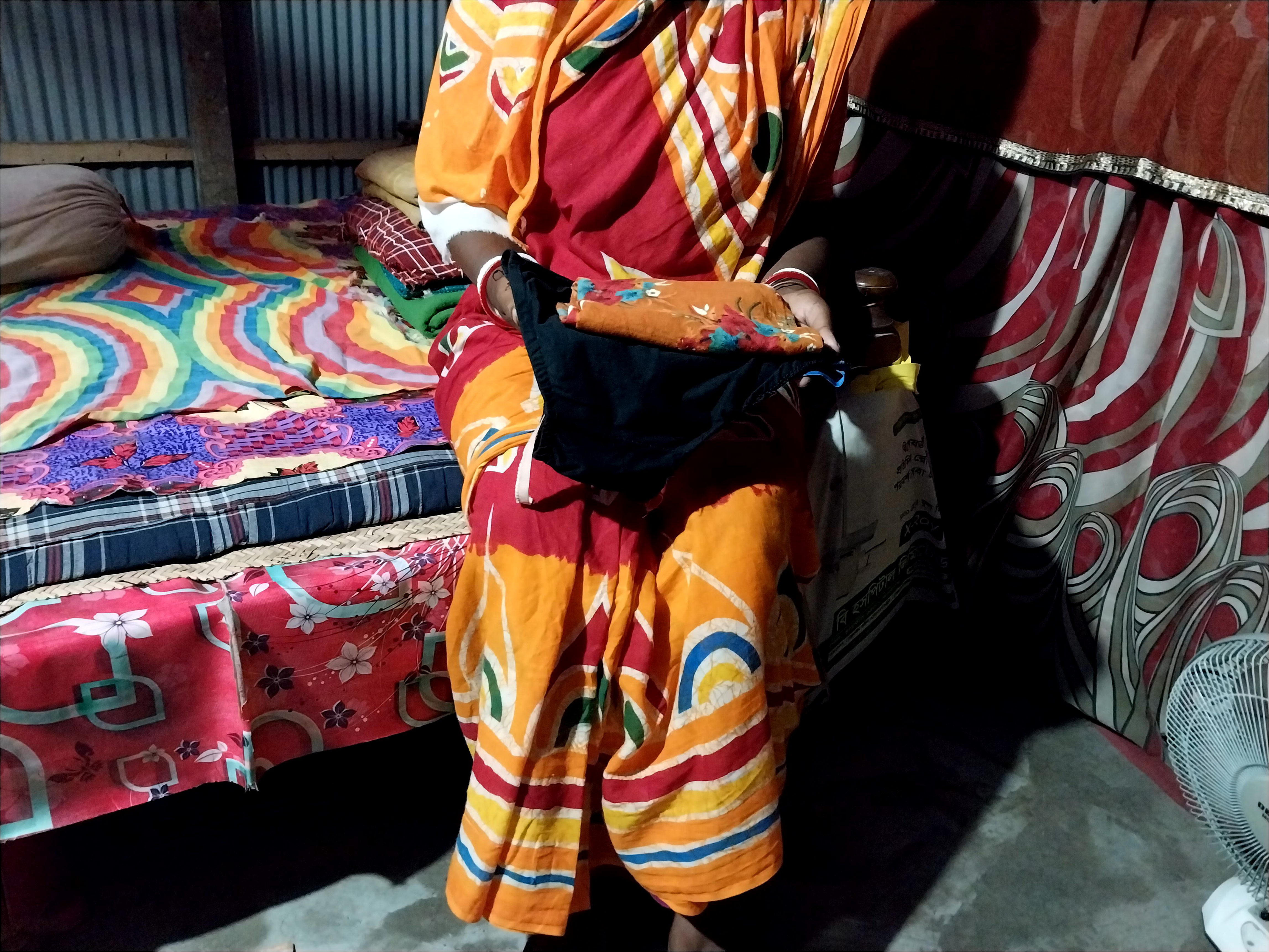
“I could not go outside in the cyclone to wash those clothes. So, I kept those used clothes in the room and the cloth used to smell.” ( Caregiver, Satkhira).

Floods in Gaibandha worsened access to clean water for washing menstrual materials, forcing people to use contaminated floodwater. One caregiver described the situation: *“During the flood, there was flood water everywhere…. our tube well was also flooded… I had to wash her menstrual clothes in flood water.” (Caregiver, Gaibandha)*

Typically, participants and their caregivers dried menstrual materials in direct sunlight outside, indicating positive menstrual behaviours: *“After she uses the cloth…I wash her cloth and dry it in the sun, so no germ remains in the cloth.”* (Female with disability, Satkhira)

However, adverse weather conditions during cyclones and floods made outdoor drying unfeasible for most participants in both districts. Therefore, they had to dry their menstrual cloths inside the home, as reported by the caregiver of a female with disability who was affected by the flood:

“*Usually, she puts the cloths on a rope in the sunlight. But during the flood, she put it (the cloth) to dry in the corner of our house…but there was no sunlight in that place…”* (Caregiver, Gaibandha)

No participants had disposal systems for their menstrual materials, relying on burying worn-out menstrual cloths near their houses. However, due to the climate hazards, these practices were impossible. For instance, one of the caregivers from Satkhira reported that instead of burying her daughter’s menstrual cloth, she stored them at home until the storm dissipated.

“*I buried the cloths under the ground after she used them during normal days… I could not dispose of them at the time of the cyclone. I kept those in the corner of the room, wrapped in paper. Then, when the storm/rain stopped, I buried them outside under the ground*.” (Caregiver, Satkhira)

While others continued to wash and reuse during the weather event, two participants from Gaibandha needed to dispose of their worn-out menstrual cloths, so they threw them in the floodwater surrounding their homes.

*“I threw them (the cloths) into the flood water because at that time I could not go to the backyard…it was very difficult for me to pass through the flood water.”* (Female with disability, Gaibandha)

The inability to maintain menstrual health during climate hazards negatively affected their health and well-being. Two women with disabilities who experienced incontinence and menstruation during climate hazards had rashes, allergies, or challenges washing clothes frequently enough. *“I changed her [menstrual] cloths less frequently at that time… Due to remaining in unclean condition for a long time, she faced allergies and rashes in her private part due to waste(faecal) and menstruation.”* (Caregiver, Satkhira)

*“She defecates several times on her clothes, and she doesn’t understand when it’s time to defecate….For her menstruation, we cut the old used clothes for her. During the flood, I washed those in the flood water. She needed to change the cloth many times because she ruined her cloths several times a day.”* (Caregiver, Gaibandha)

Two women with disabilities who evacuated their homes with their caregivers during the floods were menstruating. Both reported a reduced sense of privacy and struggled with changing, cleaning, and drying their menstrual materials in a makeshift shelter, which was not a designated evacuation shelter.

“*At that time, I changed my menstrual cloths and cleaned my private parts in our roadside makeshift house…I felt shy and nervous… I thought people could see me when I changed my menstrual cloths. I felt shy*.” (Female with disability, Gaibandha)

*“Because we were staying at the NGO office during the flood… many men and women were there. There was no place to dry the [menstrual] cloths…People could see those.”* (Caregiver, Gaibandha)

Moreover, caregivers faced the emotional strain of balancing support for the person’s menstruation with other caregiving tasks, which intensified during cyclones and floods. *“During the flood, I had to wash her menstrual clothes in flood water. It was tough for me…. Sometimes I feel irritated. But I cannot show her my feelings. If I show her that I am bothered, she will feel hurt…My life is miserable…” (*Caregiver, Gaibandha)

Tellingly, a caregiver from Satkhira expressed relief that her daughter with multiple impairments did not menstruate during the cyclone since it would have increased her hygiene-related tasks.

*“No, no, she was not menstruating during the cyclone; it was a relief. We were saved a big time. If she had been menstruating during the cyclone, it would have raised more problems. If she had been menstruating then, I [would have] had to clean everything.”* (Caregiver, Satkhira)

## Discussion

This study examined how climate hazards impacted the personal hygiene experiences of people with disabilities and their caregivers in Satkhira and Gaibandha districts, Bangladesh, focusing on their management strategies. Cyclone Amphan (2020) and recurrent severe flooding (2019– 2023) in these districts created significant challenges, including limited access to bathing and laundry facilities, water points, pathways, and submerged tube wells. Strategies to meet hygiene needs included avoiding bathing and laundry or using contaminated floodwaters. Managing incontinence and menstrual health proved especially difficult, leading to skin infections, emotional distress and compromised dignity, privacy, and emotional well-being. These findings underline the need for climate-resilient WASH interventions tailored to the hygiene needs of people with disabilities during climate hazards.

### Handwashing

Hand washing can be challenging for people with self-care limitations, and non-inclusive facility designs make independent access even more difficult [75–77]. While caregivers often assist, inadequate water and hygiene services worsen these challenges [53]. In both Satkhira and Gaibandha, most people with disabilities relied on their caregivers for handwashing, even under non-climate hazard conditions. Our study found that people with disabilities continued relying on their caregivers to provide water for handwashing during and after cyclones and floods, as access to water and hygiene materials remained limited. A nationwide population-based survey in Bangladesh similarly found that more people with disabilities required assistance from their caregivers to wash their hands than those without disabilities, highlighting persistent barriers to access to handwashing facilities and hygiene products [63].

### Bathing and laundry

Increased water availability supports hygiene practices by enabling bathing, laundry and cleaning [25, 49]. Stein et al. noted that climate-induced water insecurity disproportionately affects people with disabilities due to inaccessible WASH facilities, discrimination, stigma and economic barriers, particularly in low-income countries where climate hazards such as floods and droughts exacerbate water insecurity [78]. Similarly, our findings from Satkhira and Gaibandha revealed inaccessible water points and muddy, slippery and flooded paths during and after cyclones and floods made it difficult for people with disabilities and caregivers to collect and store safe water, resulting in days without bathing or laundry.

Studies from Vanuatu, Cambodia, Bangladesh, and Malawi highlight similar challenges, such as independent access and ease of using bathing and laundry facilities when needed [28, 52, 57, 79, 80]. In Malawi, people with disabilities could not collect sufficient water or access stored water for bathing and laundry due to physical difficulty, inaccessible water points, and safety concerns, resulting in dependence on caregivers for water collection [52]. Our findings expand on this by revealing how climate hazards intensify these struggles, disrupting bathing and laundry. Mactaggart et al.’s studies from Bangladesh, India, and Malawi found that physical limitations and disability-related stigma hindered water access [51]. While in Guatemala, people with disabilities were less likely to be able to use the same bathing facilities as others and were more likely to come in contact with dirt or contaminated water [50]. However, our findings did not identify disability-related stigma as a barrier; instead, adverse weather and damaged paths to tube wells were the primary obstacles. These differences may be attributed to our specific focus on personal hygiene in the context of climate hazards, where challenges in accessing water points are likely more pronounced. In contrast, Mactaggart et al.[51] and Kuper et al. [50] did not account for this variable. Our findings stress the need for improved water access during climate emergencies for people with disabilities to support effective hygiene behaviours and prevent adverse health effects. Accessible WASH infrastructure (for example, raised water points, bathing chairs, guide ropes, and hand and foot-operated washing facilities) constructed using universal design principles should be promoted to support safe and independent bathing and laundry.

People with disabilities and their caregivers expressed frustration over their inability to bathe regularly, as personal hygiene holds significant cultural and religious importance in Bangladesh [81]. In Satkhira, participants reported feeling unclean and dissatisfied when unable to bathe, underscoring the connection between hygiene and well-being. In flood-prone Gaibandha, many resorted to using floodwater for bathing and laundry due to submerged water sources, resulting in skin problems and fever. This aligns with other studies, such as Jerin et al., who found flood-related health complications among 280 households of people with disabilities in Jamalpur, including fever, diarrhoea, and skin diseases related to bathing and using floodwater [82].

Using floodwater poses serious health risks, particularly for people with disabilities who face heightened vulnerability due to pre-existing health conditions, poverty, stigma, discrimination, limited healthcare access, inadequate understanding by healthcare workers and lack of social support [83]. Floodwater, which is often contaminated with faeces, carries pathogens that increase the risk of infectious diseases such as cholera, diarrhoea, dysentery, rotavirus, typhoid, and intestinal parasitic diseases like amoebiasis and cryptosporidiosis [84–89]. This suggests that people with disabilities are at greater risk of experiencing disproportionately negative health outcomes when forced to use contaminated floodwater for bathing and laundry. Their pre-existing healthcare challenges are significantly worsened during such emergencies, making them even more vulnerable. Therefore, clean water should be ensured to enable frequent handwashing, bathing, and doing laundry during and after climate events, minimising health risks from exposure to contaminated floodwaters. Clean water supplies, such as piped or supplementary water systems, in the households of people with disabilities should be prioritised to fulfil bathing and laundry needs during and after climate events, minimising health risks from exposure to contaminated floodwaters and strengthening hygiene resilience against climate hazards.

### Managing incontinence

Managing hygiene for individuals with incontinence requires significantly more water and soap for washing than for those without incontinence [27]. Studies from Bangladesh, Ethiopia, Ghana, Malawi, and Vanuatu highlight increased demands for water, hygiene products, and accessible bathing and laundry facilities for incontinence management [26–28, 53]. In Vanuatu, Mactaggart et al. found that people with disabilities experiencing incontinence often remained soiled overnight due to inaccessible latrines [30], leading to health risks from prolonged exposure to excreta.

In Satkhira, our study found that individuals with incontinence and their caregivers faced severe challenges during cyclones, as they could not reach bathing facilities despite needing immediate washing after urinating or defecating. Some remained soiled for extended periods until the cyclone subsided, compromising the support caregivers could provide with washing and cleaning and affecting recipients’ privacy and dignity. In Gaibandha, caregivers resorted to washing soiled clothes in floodwater, increasing infection risks. Poor hygiene for those living with incontinence can lead to dermatitis, skin infections, diarrhoeal diseases, and urinary tract infections [27]. These findings suggest that inadequate hygiene for people with disabilities experiencing incontinence during climate hazards significantly worsens health risks, highlighting an urgent need for targeted interventions. Therefore, affordable incontinence products such as absorbent pads, commodes, and hygiene products should be distributed alongside guidance for caregivers on managing soiling amidst climate hazards and ensuring dignity during bathing and laundry.

### Menstrual health

Our findings indicate that women with disabilities and their caregivers knew the importance of washing menstrual materials with soap and clean water. However, the cyclone in Satkhira hindered these practices due to adverse weather conditions, which made it nearly impossible to leave the house to collect water and resulted in a shortage of clean water for washing menstrual materials. These findings align with other humanitarian contexts where limited water resources affected the ability of women with disabilities to maintain menstrual health, including washing materials and cleaning their bodies [90]. For example, due to water shortages, Nuzhat et al. found that cyclone-affected women from coastal Bangladesh used saline water to wash their bodies and menstrual materials [91]. Still, studies have not yet focused on how these challenges specifically impact women with disabilities. This underscores the urgent need for further research into how water inaccessibility during climate hazards affects menstrual health.

Existing evidence indicates that lack of safe water can contribute to feelings of powerlessness, fear, and depression among people with disabilities. This is coupled with the mental stresses of surviving climate hazards involving evacuation and relocation during floods and cyclones [92–94]. During the floods in Gaibandha, women with disabilities who menstruated and their caregivers resorted to using floodwater to wash menstrual materials due to the submerged tube wells and surroundings. Similar strategies have been reported from flood-prone Jamalpur, where women used polluted floodwaters to wash their menstrual materials [82]. Previous studies also highlight the health risks of using poor-quality water during menstruation (e.g., floodwater or saline waters), such as discomfort, rashes, burns, and urinary tract infections [91].

The use of poor-quality water for washing menstrual materials exacerbates vulnerabilities, especially for women with disabilities, underscoring the need for safe water access to protect health and dignity during such emergencies.

Women with disabilities and their caregivers in both districts also struggled with drying menstrual materials because of heavy rain, lack of sunlight and stagnant floodwater. Other studies from Bangladesh [91] and Vanuatu echo this [95]. Furthermore, incomplete drying of reusable menstrual cloths can lead to perennial rash and urinary tract infections, as evidenced in other contexts [8, 96]. Disposal was equally challenging, as many women could not bury or discard used materials due to damaged paths and heavy rain, often keeping them inside the house. Safe, hygienic and environmentally friendly disposal mechanisms are lacking in many settings, which risks further environmental degradation [60, 97–100]. For example, women with and without disabilities often dispose of menstrual products inappropriately, such as in rivers and down hillsides, due to a lack of private disposal options and awareness of environmental impacts [59, 60, 98, 100]. This practice is particularly prevalent in LMICs and increases the risks of social and psychological distress [60, 63, 98–100]. This poses risks of disease transmission, environmental harm, and stress, emphasising the urgent need for accessible disposal facilities and infrastructure during climate hazards [99, 101].

Privacy and safety concerns further complicated menstrual health for women with disabilities in temporary shelters during floods. In flood-prone Gaibandha, women sought refuge in various places, including temporary shelters. In these conditions, their privacy was often compromised. Similar challenges related to privacy, comfort and safety for menstrual health have been reported in other settings. For instance, in Vanuatu, women with disabilities who were menstruating often waited until nightfall to bathe in relative privacy [57]. Caregivers of daughters with intellectual disabilities who had evacuated to cyclone shelters in Vanuatu reported returning to their damaged households to wash and change their daughters’ menstrual materials behind the remaining banana trees, as this provided greater privacy and safety than the cyclone shelters [102]. In Pakistan, displaced women struggled with the absence of menstrual materials and suitable private places for changing and disposal, while in India, the absence of separate toilets and proper disposal systems exacerbated these issues for displaced women [103, 104]. Improving privacy, safety, and sanitation in shelters is essential to support menstrual health for women with disabilities and their caregivers during climate crises.

Our study found that no woman with a disability or their caregiver evacuated their homes during the cyclone in Satkhira and took shelter in the designated cyclone shelters. Only two women with disabilities and their caregivers who were menstruating during the floods in Gaibandha evacuated their homes. Both cited privacy concerns regarding maintaining menstrual health in temporary shelters. However, it is difficult to conclude that women with disabilities do not evacuate their homes during climate hazards in Bangladesh from the experiences of only two participants. Yet, other studies do suggest this can be a challenge. Women with disabilities in Vanuatu reported avoiding evacuation during menstruation due to privacy concerns, fear of gender-based violence, and anxiety from potential menstrual leakage [95]. Further research is needed to fully understand the specific challenges and develop evidence-based options when using temporary shelters faced by women with disabilities across settings.

Although only two women with disabilities were found to experience both incontinence and menstruation during climate hazards, their experiences highlight unique challenges. In Satkhira, one participant reported changing menstrual and incontinence materials less frequently due to difficulties with washing, drying and disposal during a cyclone, which could lead to rashes, itching and infections [105]. Conversely, in Gaibandha, another participant frequently washed her menstrual cloth and soiled clothes using floodwater due to a lack of safe water. These findings underscore the need for safe water, accessible and affordable incontinence and menstrual products, and accessible spaces for washing and drying materials to minimise health risks during climate hazards. The findings also highlight that disaster preparedness plans should include accessible hygiene kits containing soap, menstrual and incontinence materials, drying and disposal options, and training for caregivers to manage menstrual health and incontinence safely, hygienically and with dignity during climate hazards.

Our study emphasises the connection between disability, disrupted hygiene practices due to climate hazards, and adverse health effects, such as fever, skin problems, menstrual health issues, and overall well-being. Moreover, these disruptions lead to increased dependency on caregivers, affecting their emotional well-being. Caregivers often struggle to address hygiene needs, particularly in the absence of adequate support, increasing their levels of stress and vulnerability. Integrating disability-inclusive approaches into WASH-related adaptation and mitigation strategies is critical to address these issues. This includes ensuring adequate access to safe water, accessible hygiene facilities, and sufficient hygiene materials during disaster preparedness, response and recovery efforts. Co-developing emergency preparedness tools with people with disabilities and caregivers, promoting safe and dignified hygiene practices through behaviour change campaigns, and ensuring climate-resilient, safe, and private WASH facilities in households and temporary shelters will complement these efforts. Inclusive planning must prioritise context-specific interventions and actively engage people with disabilities and their caregivers through consultation in designing solutions that strengthen resilience to recurring climate hazards. Expanding research to diverse geographic regions and climate hazard scenarios can provide broader insights and inform strategies for inclusive WASH services.

### Strengths and limitations

One strength of this study is its in-depth exploration of the unique challenges faced by people with disabilities during climate hazards in Bangladesh, a topic often underrepresented in disaster and public health research. The qualitative approach, using various data collection tools, provided a comprehensive understanding of personal experiences and the strategies employed by individuals with disabilities and their caregivers to manage personal hygiene. Secondly, the study covers a wide range of hygiene practices of people with disabilities, including an emphasis on persons experiencing incontinence, including bathing, laundry, and menstrual health, offering a holistic view of personal hygiene experiences.

Regarding study limitations, we did not include epidemiological evidence linking inadequate hygiene practices to specific health outcomes, as this was beyond the scope of our research. Therefore, any association between disease and hygiene is anecdotal. Yet, this remains a critical area for future investigation. Additionally, the findings may not be generalisable for other climate hazards, such as reduced rainfall and drought. The study’s focus on the immediate aftermath of climate hazards may also overlook long-term effects on hygiene practices and health outcomes in the changing climate.

## Conclusion

This study highlights the severe challenges faced by people with disabilities and their caregivers in Bangladesh in maintaining personal hygiene - including bathing, laundry, handwashing, incontinence management, and menstrual health - during and after climate hazards such as cyclones and floods. Climate hazards disrupt access to clean WASH facilities, forcing reliance on contaminated floodwater or cessation of hygiene practices. These disruptions lead to dissatisfaction, compromised privacy and dignity, and increased health risks—particularly for individuals managing incontinence or those residing in temporary shelters or living in isolated and waterlogged areas.

This study offers valuable insights into the intersection of disability, personal hygiene, and climate hazards in Bangladesh. This underscores the critical need for disability-inclusive, climate-resilient WASH services, interventions, and policies. By documenting the experiences of people with disabilities and the health impacts of inadequate hygiene during climate hazards, our study highlights the importance of targeted measures to safeguard the health, dignity, and resilience of people with disabilities and their caregivers. Actively involving people with disabilities in designing and implementing interventions would enhance community resilience and promote inclusive hygiene practices amid increasing climate challenges. Urgent resources are needed to address these issues and prevent further intensification of inequalities for people with disabilities.

## Supporting information

Guideline to interview people with disabilities

Guideline to interview caregivers

Guideline for accessibility and safety audit with people with disabilities

Photovoice and ranking guideline

## Data Availability

All relevant data are within the manuscript and its Supporting Information files.

## Acknowledgements

We extend our gratitude to all the study participants who openly shared their experiences for this study. We also acknowledge the contributions of Naila Ferdousi Haque and Sabiha Ahmed Diba to the broader research on how human-induced climate change affects the WASH experiences of people with disabilities in Bangladesh. We also thank the World Vision and Organisations of Persons with Disabilities (OPDs) from Satkhira and Gaibandha for their insights regarding disaster management and hygiene and for their support in communicating with people with disabilities.

## Authors contribution

Conceptualisation: Jane Wilbur, Mahbub-Ul Alam

Data Curation: Shahpara Nawaz, Bithy Podder, Arka Goshami, Tasnia Alam Upoma Formal Analysis: Shahpara Nawaz, Tasnia Alam Upoma, Bithy Podder, Arka Goshami, Funding acquisition: Jane Wilbur, Mahbub-Ul Alam

Investigation: Shahpara Nawaz, Jane Wilbur, Mahbub-Ul Alam, Bithy Podder, Arka Goshami, Mehedi Hasan

Methodology: Jane Wilbur, Mahbub-Ul Alam, Shahpara Nawaz, Tasnia Alam Upoma Project Administration: Shahpara Nawaz, Jane Wilbur, Mahbub-Ul Alam Supervision: Jane Wilbur, Mahbub-Ul Alam

Visualization: Shahpara Nawaz, Jarin Akter

Writing - Original draft preparation: Shahpara Nawaz, Tasnia Alam Upoma

Writing – review and editing: Shahpara Nawaz, Jane Wilbur, Tasnia Alam Upoma, Lauren D’Mello-Guyett, Jarin Akter, Mehedi Hasan, Dewan Muhammad Shoaib, Sari Kovats, Mahbub- Ul Alam

## Supporting information captions

**S1.** Guideline to interview people with disabilities

**S2.** Guideline to interview caregivers

**S3.** Guideline for accessibility and safety audit with people with disabilities

**S4.** Photovoice and ranking guideline

## References

1. Khatoon R, Sachan B, Khan MA, Srivastava JP. Impact of school health education program on personal hygiene among school children of Lucknow district. Journal of Family Medicine and Primary Care. 2017;6(1):97–100.

2. WHO, UNICEF. Hygiene 2021 [Available from: https://washdata.org/monitoring/hygiene#:~:text=Hygiene%20refers%20to%20the%20conditions,management%20(see%20Menstrual%20Health).

3. Chakraborty TK, Kabir A, Ghosh GC. Impact and adaptation to cyclone AILA: focus on water supply, sanitation and health of rural coastal community in the south west coastal region of Bangladesh. Journal of Health and Environmental Research. 2016;2(3):13–9.

4. Curtis V, Cairncross S, Yonli R. Review: Domestic hygiene and diarrhoea – pinpointing the problem. Tropical Medicine & International Health. 2000;5(1):22–32.

5. Haque MA, Haque A, Ansari M. Water, sanitation and health status of Aila affected coastal area of Bangladesh. Bangladesh J Environ Sci. 2010;19:51–6.

6. Oloruntoba EO, Folarin TB, Ayede AI. Hygiene and sanitation risk factors of diarrhoeal disease among under-five children in Ibadan, Nigeria. African health sciences. 2014;14(4):1001–11.

7. Prüss-Ustün A, Wolf J, Bartram J, Clasen T, Cumming O, Freeman MC, et al. Burden of disease from inadequate water, sanitation and hygiene for selected adverse health outcomes: An updated analysis with a focus on low- and middle-income countries. International Journal of Hygiene and Environmental Health. 2019;222(5):765–77.

8. Al Karmi J, Alshrouf MA, Haddad TA, Alhanbali AE, Raiq NA, Ghanem H, et al. Urinary and reproductive tract infection symptoms and menstrual hygiene practices in refugee camps in Jordan: A cross-sectional study. Women’s Health. 2024;20:17455057241240920.

9. Baker KK, Padhi B, Torondel B, Das P, Dutta A, Sahoo KC, et al. From menarche to menopause: A population-based assessment of water, sanitation, and hygiene risk factors for reproductive tract infection symptoms over life stages in rural girls and women in India. PLOS ONE. 2017;12(12):e0188234.

10. Das P, Baker KK, Dutta A, Swain T, Sahoo S, Das BS, et al. Menstrual Hygiene Practices, WASH Access and the Risk of Urogenital Infection in Women from Odisha, India. PLOS ONE. 2015;10(6):e0130777.

11. Nabwera HM, Shah V, Neville R, Sosseh F, Saidykhan M, Faal F, et al. Menstrual hygiene management practices and associated health outcomes among school-going adolescents in rural Gambia. PLOS ONE. 2021;16(2):e0247554.

12. Campbell SJ, Biritwum N-K, Woods G, Velleman Y, Fleming F, Stothard JR. Tailoring Water, Sanitation, and Hygiene (WASH) Targets for Soil-Transmitted Helminthiasis and Schistosomiasis Control. Trends in Parasitology. 2018;34(1):53–63.

13. Freeman MC, Chard AN, Nikolay B, Garn JV, Okoyo C, Kihara J, et al. Associations between school- and household-level water, sanitation and hygiene conditions and soil- transmitted helminth infection among Kenyan school children. Parasites & Vectors. 2015;8(1):412.

14. Mohammadpour M, Abrishami M, Masoumi A, Hashemi H. Trachoma: Past, present and future. Journal of Current Ophthalmology. 2016;28(4):165–9.

15. Birawida AB, Mallongi A, Satrianegara FM, Khaer A, Appolo A, Restu M. Factors Related to the Incidence of Contact Dermatitis In-Fisherman on the Spermonde Island. Open Access Macedonian Journal of Medical Sciences. 2020;8(T2):220–3.

16. Haradanhalli RS, Prashanth RM, Kumari N, Siddhareddy I, Surendran J. Personal hygiene practices and related skin diseases among primary school children of urban poor locality. International Journal of Community Medicine and Public Health. 2019;6(6):2526.

17. Al-Rifaai JM, Al Haddad AM, Qasem JA. Personal hygiene among college students in Kuwait: A Health promotion perspective. Journal of Education and Health Promotion. 2018;7(1):92.

18. Bisung E, Elliott SJ. Psychosocial impacts of the lack of access to water and sanitation in low- and middle-income countries: a scoping review. Journal of Water and Health. 2016;15(1):17–30.

19. Hutton G, Chase C. Water supply, sanitation, and hygiene. In: Mock CN, Nugent R KO, editors. Injury Prevention and Environmental Health. 3: The World Bank; 2018.

20. Ross I, Bick S, Ayieko P, Dreibelbis R, Wolf J, Freeman MC, et al. Effectiveness of handwashing with soap for preventing acute respiratory infections in low-income and middle- income countries: a systematic review and meta-analysis. The Lancet. 2023;401(10389):1681–90.

21. Wolf J, Johnston RB, Ambelu A, Arnold BF, Bain R, Brauer M, et al. Burden of disease attributable to unsafe drinking water, sanitation, and hygiene in domestic settings: a global analysis for selected adverse health outcomes. Lancet. 2023;401(10393):2060–71.

22. CDC. Water, Sanitation, and Environmentally Related Hygiene (WASH). 2024. Available from: https://www.cdc.gov/hygiene/about/index.html.

23. Hennegan J, Winkler I, Bobel C, Keiser J, Hampton J, Larsson G, et al. Menstrual health: a definition for policy, practice, and research. Sexual and Reproductive Health Matters. 2021;29.

24. Mactaggart I, Baker S, Bambery L, Iakavai J, Kim MJ, Morrison C, et al. Water, women and disability: Using mixed-methods to support inclusive WASH programme design in Vanuatu. The Lancet Regional Health – Western Pacific. 2021;8.

25. González Hernández L, Romano A, Hamid DM, Elsimat EAA, Ongara D, Yassin Y, et al. Left alone and behind: Experiences of living with incontinence in a Sudanese refugee camp and how WASH practitioners can support. Journal of Water, Sanitation and Hygiene for Development. 2024;14(7):521–31.

26. Rosato-Scott C, Adjorlolo S, Farrington M, Barrington DJ. ‘Do not forget us’: the shared experiences and needs of people living with incontinence in humanitarian contexts. Journal of Water, Sanitation and Hygiene for Development. 2024;14(3):220–8.

27. Rosato-Scott C, Barrington DJ, Bhakta A, House SJ, Mactaggart I, Wilbur J. Incontinence: we need to talk about leaks. Brighton, IDS; 2020. Available from: https://researchonline.lshtm.ac.uk/id/eprint/4664694/.

28. Wilbur J, Morrison C, Bambery L, Tanguay J, Baker S, Sheppard P, et al. “I’m scared to talk about it”: exploring experiences of incontinence for people with and without disabilities in Vanuatu, using mixed methods. The Lancet Regional Health–Western Pacific. 2021;14.

29. Collins SM, Mbullo Owuor P, Miller JD, Boateng GO, Wekesa P, Onono M, et al. ’I know how stressful it is to lack water!’ Exploring the lived experiences of household water insecurity among pregnant and postpartum women in western Kenya. Glob Public Health. 2019;14(5):649–62.

30. Mactaggart I, Baker S, Bambery L, Iakavai J, Kim MJ, Morrison C, et al. Water, women and disability: Using mixed-methods to support inclusive wash programme design in Vanuatu. The Lancet Regional Health–Western Pacific. 2021;8.

31. Wilbur J, Ruuska D, Nawaz S, Natukunda J. Climate Risks to Water, Sanitation and Hygiene Services and Evidence of Inclusive and Effective Interventions in Low and Middle- Income Countries: A Scoping Review. medRxiv. 2024:2024.08.21.24312122.

32. Centre for Climate Change and Environmental Research (C3ER) - BRAC University, Centre for Disability and Development, CBM. Study on Disability Inclusive Climate Change Adaptation (DiCCA), Final report. Available at https://cdd.org.bd/wp-content/uploads/2022/05/Study-Report.pdf (accessed 18 February 2023); 2022.

33. Andrade L, O’Dwyer J, O’Neill E, Hynds P. Surface water flooding, groundwater contamination, and enteric disease in developed countries: A scoping review of connections and consequences. Environmental Pollution. 2018;236:540–9.

34. Dura G, Pándics T, Kádár M, Krisztalovics K, Kiss Z, Bodnár J, et al. Environmental health aspects of drinking water-borne outbreak due to karst flooding: case study. Journal of Water and Health. 2010;8(3):513–20.

35. Fredrick T, Ponnaiah M, Murhekar MV, Jayaraman Y, David JK, Vadivoo S, et al. Cholera outbreak linked with lack of safe water supply following a tropical cyclone in Pondicherry, India, 2012. J Health Popul Nutr. 2015;33(1):31-8.

36. McCordic C, Raimundo I, Judyn M, Willis D. The distribution of Cyclone Idai’s water impacts in Beira, Mozambique. Disaster Prevention and Management: An International Journal. 2024;33(6):1–15.

37. Rafa N, Jubayer A, Uddin SMN. Impact of cyclone Amphan on the water, sanitation, hygiene, and health (WASH2) facilities of coastal Bangladesh. Journal of Water, Sanitation and Hygiene for Development. 2021;11(2):304–13.

38. Bhat VN. Impact of droughts on industrial emissions into surface waters. Environmental Science and Pollution Research. 2020;27(34):42806–14.

39. Graham DJ, Bierkens MFP, van Vliet MTH. Impacts of droughts and heatwaves on river water quality worldwide. Journal of Hydrology. 2024;629:130590.

40. Hrdinka T, Novický O, Hanslík E, Rieder M. Possible impacts of floods and droughts on water quality. Journal of Hydro-environment Research. 2012;6(2):145–50.

41. Veijalainen N, Ahopelto L, Marttunen M, Jääskeläinen J, Britschgi R, Orvomaa M, et al. Severe Drought in Finland: Modeling Effects on Water Resources and Assessing Climate Change Impacts. Sustainability. 2019;11(8):2450.

42. Zamrsky D, Oude Essink GHP, Bierkens MFP. Global Impact of Sea Level Rise on Coastal Fresh Groundwater Resources. Earth’s Future. 2024;12(1):e2023EF003581.

43. Zelenakova M, Abd-Elhamid H, Barańczuk K, Barańczuk J, editors. Assessment and adaptation to climate change and sea level rise impacts on Egypt’s northern coasts. IOP Conference Series: Materials Science and Engineering; 2022: IOP Publishing.

44. Moore E. The Effects of Climate Change on the Menstrual Health of Women and Girls in Rural Settings within Low-Income Countries: Mailman School of Public Health, Columbia University; 2022.

45. Dembedza VP, Chopera P, Macheka L. Water, Sanitation and Hygiene practices in areas affected by Cyclone Idai in Zimbabwe. Journal of Water, Sanitation and Hygiene for Development. 2024;14(7):532–42.

46. Siddik MA, Islam ARMT. A systematic review of cyclonic disaster: Damage-loss, consequences, adaptation strategies, and future scopes. Heliyon. 2024;10(12).

47. Local Government Division. National Strategy for Water Supply and Sanitation 2014. In: Ministry of Local Government RDaC, editor. Dhaka, Bangladesh: Ministry of Local Government, Rural Development and Cooperatives,Government of the People’s Republic of Bangladesh; 2014.

48. Oettle NM, Koelle BR, Schmiedel U, Archer E. Participatory Adaptation Handbook: a practitioner’s guide for facilitating people centred adaptation to climate change. Available at https://www.socialscienceinaction.org/resources/a-practitioners-guide-for-facilitating-people-centred-adaptation-participatory-adaptation-handbook/ 2014.

49. World Health Organization. Addressing climate change: supplement to the WHO water, sanitation and hygiene strategy 2018–2025: World Health Organization; 2023.

50. Kuper H, Mactaggart I, White S, Dionicio C, Cañas R, Naber J, et al. Exploring the links between water, sanitation and hygiene and disability; Results from a case-control study in Guatemala. PLOS ONE. 2018;13(6):e0197360.

51. Mactaggart I, Schmidt W-P, Bostoen K, Chunga J, Danquah L, Halder AK, et al. Access to water and sanitation among people with disabilities: results from cross-sectional surveys in Bangladesh, Cameroon, India and Malawi. BMJ Open. 2018;8(6):e020077.

52. White S, Kuper H, Itimu-Phiri A, Holm R, Biran A. A Qualitative Study of Barriers to Accessing Water, Sanitation and Hygiene for Disabled People in Malawi. PLOS ONE. 2016;11(5):e0155043.

53. Wilbur J, Dreibelbis R, Mactaggart I. Addressing water, sanitation and hygiene inequalities: A review of evidence, gaps, and recommendations for disability-inclusive WASH by 2030. PLOS Water. 2024;3(6):e0000257.

54. Ding Z, Zheng H, Wang J, O’Connor P, Li C, Chen X, et al. Integrating Top-Down and Bottom-Up Approaches Improves Practicality and Efficiency of Large-Scale Ecological Restoration Planning: Insights from a Social–Ecological System. Engineering. 2022.

55. Ahmed S, Hasan MZ, Pongsiri MJ, Ahmed MW, Szabo S. Effect of extreme weather events on injury, disability, and death in Bangladesh. Climate and Development. 2021;13(4):306–17.

56. Lindsay S, Hsu S, Ragunathan S, Lindsay J. The impact of climate change related extreme weather events on people with pre-existing disabilities and chronic conditions: a scoping review. Disability and Rehabilitation. 2023;45(25):4338–58.

57. Wilbur J, Morrison C, Iakavai J, Shem J, Poilapa R, Bambery L, et al. “The weather is not good”: exploring the menstrual health experiences of menstruators with and without disabilities in Vanuatu. The Lancet Regional Health–Western Pacific. 2022;18.

58. Chowdhury MA, Nowreen S, Tarin NJ, Hasan MR, Zzaman RU, Amatullah NI. WASH and MHM experiences of disabled females living in Dhaka slums of Bangladesh. Journal of Water, Sanitation and Hygiene for Development. 2022;12(10):683–97.

59. Wilbur J, Torondel B, Hameed S, Mahon T, Kuper H. Systematic review of menstrual hygiene management requirements, its barriers and strategies for disabled people. PLOS ONE. 2019;14(2):e0210974.

60. Wilbur J, Kayastha S, Mahon T, Torondel B, Hameed S, Sigdel A, et al. Qualitative study exploring the barriers to menstrual hygiene management faced by adolescents and young people with a disability, and their carers in the Kavrepalanchok district, Nepal. BMC Public Health. 2021;21(1):476.

61. Hashizume M, Wagatsuma Y, Faruque AS, Hayashi T, Hunter PR, Armstrong B, et al. Factors determining vulnerability to diarrhoea during and after severe floods in Bangladesh. Journal of water and health. 2008;6(3):323–32.

62. Jimenez MP, DeVille NV, Elliott EG, Schiff JE, Wilt GE, Hart JE, et al. Associations between nature exposure and health: a review of the evidence. International journal of environmental research and public health. 2021;18(9):4790.

63. Alam MU, Tabassum KF, M. SD, Hasan M, Upoma TA, Rahat A, et al. Assessment of WASH and incontinence situation of people with disabilities and older people in Bangladesh. London; 2023. Available from: https://www.lshtm.ac.uk/sites/default/files/2023-11/Assessment%20of%20WASH%20and%20incontinence%20situation%20of%20people%20with%20disabilities%20and%20older%20people%20in%20Bangladesh.pdf?ref=disabilitydebrief.org.

64. 64. WaterAid Bangladesh. Climate Resilient WASH Programming in Coastal Areas of Bangladesh. An end-line study. Available at https://www.wateraid.org/bd/sites/g/files/jkxoof236/files/climate-resilience-wash-programming-in-coastal-areas-of-bangladesh-an-end-line-study_0.pdf (accessed 18 February 2023); No date.

65. Zhaowei D, Hua Z, Jun W, Patrick OC, Cong L, Xiaodong C, et al. Integrating Top-Down and Bottom-Up Approaches Improves Practicality and Efficiency of Large-Scale Ecological Restoration Planning: Insights from a Social-Ecological System. Engineering. 2022.

66. Ministry of Environment Forest and Climate Change, Government of Bangladesh. National Adaptation Plan of Bangladesh (2023-2050). Available at https://www4.unfccc.int/sites/SubmissionsStaging/Documents/202211020942---National%20Adaptation%20Plan%20of%20Bangladesh%20(2023-2050).pdf2022.

67. Ministry of Environment and Forest. Bangladesh Climate Change Stratergy and Action Plan (BCCSAP) 2009. In: Forest MoEa, editor. Dhaka, Bangladesh: Government of the People’s Republic of Bangladesh 2009. p. 98.

68. Legislative and Parliamentary Affairs Division. Persons with disabilities rights and protection act in Bangladesh (2013). In: Government of the People’s Republic of Bangladesh, editor. Dhaka, Bangladesh: Government of the People’s Republic of Bangladesh,; 2013. p. 27.

69. Scherer N, Mactaggart I, Huggett C, Pheng P, Rahman MU, Biran A, et al. The Inclusion of Rights of People with Disabilities and Women and Girls in Water, Sanitation, and Hygiene Policy Documents and Programs of Bangladesh and Cambodia: Content Analysis Using EquiFrame. Int J Environ Res Public Health. 2021;18(10).

70. 70. NIRAPAD, Start Network, UNCT Bangladesh. Multi-Hazard Risk Analysis of Climate- Related Disasters in Bangladesh. 2021. Available from: https://reliefweb.int/report/bangladesh/multi-hazard-risk-analysis-climate-related-disasters-bangladesh.

71. Alam MZ, Carpenter-Boggs L, Mitra S, Haque MM, Halsey J, Rokonuzzaman M, et al. Effect of salinity intrusion on food crops, livestock, and fish species at Kalapara Coastal Belt in Bangladesh. Journal of Food Quality. 2017;2017.

72. Washington Group on Disability Statistics. The Washington Group Short Set on Functioning – Enhanced (WG-SS Enhanced): University College London 2022 [5]. Available from: https://www.washingtongroup-disability.com/question-sets/wg-short-set-on-functioning-enhanced-wg-ss-enhanced/.

73. Centre for Research on the Epidemiology of Disasters (CRED). EM-DAT:The International Disaster Database: University of Louvain (UCLouvain); 2023 [Available from: https://public.emdat.be/data.

74. Ghosh BK, Sarker KR. Climate Change and Living with Floods: An Empirical Case from the Saghata Union of Gaibandha District, Bangladesh. Bangladesh II: Climate Change Impacts, Mitigation and Adaptation in Developing Countries. Cham: Springer International Publishing; 2021. p. 459-78.

75. McLay L, van Deurs J, Gibbs R, Whitcombe-Dobbs S. Empirically Supported Strategies for Teaching Personal Hygiene Skills to People with Intellectual Disabilities. In: Lang R, Sturmey P, editors. Adaptive Behavior Strategies for Individuals with Intellectual and Developmental Disabilities: Evidence-Based Practices Across the Life Span. Cham: Springer International Publishing; 2021. p. 47-72.

76. Oloruntoba EO, Udofia IP, Adejumo M. Access to sanitation facilities and handwashing practices among physically challenged persons in homes for the disabled in Ibadan, Nigeria. Journal of Environmental Protection. 2020;11(04):299.

77. Wijayasingha LNS, Lo B, editors. A wearable sensing framework for improving personal and oral hygiene for people with developmental disabilities. 2016 IEEE Wireless Health (WH); 2016 25-27 Oct. 2016.

78. Stein PJS, Stein MA, Groce N, Kett M, Akyeampong EK, Alford WP, et al. Advancing disability-inclusive climate research and action, climate justice, and climate-resilient development. The Lancet Planetary Health. 2024;8(4):e242–e55.

79. Wilbur J, Pheng P, Has R, Nguon SK, Banks LM, Huggett C, et al. A qualitative cross-sectional study exploring the implementation of disability-inclusive WASH policy commitments in Svay Reing and Kampong Chhnang Provinces, Cambodia. Frontiers in Water. 2022;4.

80. Wilbur J. Translating disability-inclusive WASH policies into practice: lessons learned from Bangladesh. Translating disability-inclusive WASH policies into practice: lessons learned from Bangladesh. 2022.

81. Sultana R, Nahar N, Rimi NA, Swarna ST, Khan S, Saifullah MK, et al. The Meaning of “Hygiene” and Its Linked Practices in a Low-Income Urban Community in Bangladesh. International Journal of Environmental Research and Public Health. 2022;19(16):9823.

82. Jerin T, Chowdhury MA, Azad MAK, Zaman S, Mahmood S, Islam SLU, et al. Extreme weather events (EWEs)-Related health complications in Bangladesh: A gender-based analysis on the 2017 catastrophic floods. Natural Hazards Research. 2023.

83. Kuper H, Heydt P. The Missing Billion - Access to health services for 1 billion people with disabilities. International Centre for Evidence in Disability, London School of Hygiene and Tropical Medicine,; 2019. Available from: https://www.lshtm.ac.uk/research/centres/international-centre-evidence-disability/missing-billion.

84. Phan NK, Sherchan SP. Microbiological Assessment of Tap Water Following the 2016 Louisiana Flooding. Int J Environ Res Public Health. 2020;17(4).

85. Berahmat R, Spotin A, Ahmadpour E, Mahami-Oskouei M, Rezamand A, Aminisani N, et al. Human cryptosporidiosis in Iran: a systematic review and meta-analysis. Parasitology Research. 2017;116(4):1111–28.

86. Haghighi A, Riahi SM, Taghipour A, Spotin A, Javanian M, Mohammadi M, et al. Amoebiasis in Iran: a systematic review and meta-analysis. Epidemiology and Infection. 2018;146(15):1880–90.

87. Okaka FO, Odhiambo BDO. Relationship between Flooding and Out Break of Infectious Diseasesin Kenya: A Review of the Literature. Journal of Environmental and Public Health. 2018;2018(1):5452938.

88. Paterson DL, Wright H, Harris PNA. Health Risks of Flood Disasters. Clinical Infectious Diseases. 2018;67(9):1450–4.

89. Shokri A, Sabzevari S, Hashemi SA. Impacts of flood on health of Iranian population: Infectious diseases with an emphasis on parasitic infections. Parasite Epidemiology and Control. 2020;9:e00144.

90. Wilbur J, Clemens F, Sweet E, Banks LM, Morrison C. The inclusion of disability within efforts to address menstrual health during humanitarian emergencies: A systematised review. Frontiers in Water. 2022;4.

91. Nuzhat S, Iyer R, Qader AIA, Manzoor-Al-Islam S. A Participatory Assessment for Climate-Induced WASH Vulnerabilities in Bangladesh. Brighton: IDS: The Sanitation Learning Hub; 2023. Available from: https://sanitationlearninghub.org/resource/a-participatory-assessment-for-climate-induced-wash-vulnerabilities-in-bangladesh/.

92. Uddin T, Tasnim A, Islam MR, Islam MT, Salek AKM, Khan MM, et al. Health impacts of climate-change related natural disasters on persons with disabilities in developing countries: A literature review. The Journal of Climate Change and Health. 2024;19:100332.

93. Alexander M, Alexander J, Arora M, Slocum C, Middleton J. A bellweather for climate change and disability: educational needs of rehabilitation professionals regarding disaster management and spinal cord injuries. Spinal Cord Series and Cases. 2019;5(1):94.

94. Khine MM, Langkulsen U. The Implications of Climate Change on Health among Vulnerable Populations in South Africa: A Systematic Review. International Journal of Environmental Research and Public Health. 2023;20(4):3425.

95. Wilbur J, Poilapa R, Morrison C. Menstrual Health Experiences of People with Intellectual Disabilities and Their Caregivers during Vanuatu’s Humanitarian Responses: A Qualitative Study. International Journal of Environmental Research and Public Health. 2022;19(21):14540.

96. WHO. Gender and Health in Disasters. Geneva, Switzerland: WORLD HEALTH ORGANIZATION (WHO); 2002. Available from: https://iris.who.int/bitstream/handle/10665/68886/a85575.pdf?sequence=1&isAllowed=y.

97. Robinson HJ, Barrington DJ. Drivers of menstrual material disposal and washing practices: A systematic review. PLOS ONE. 2021;16(12):e0260472.

98. Schmitt ML, Gruer C, Clatworthy D, Kimonye C, Peter DE, Sommer M. Menstrual material maintenance, disposal, and laundering challenges among displaced girls and women in Northeast Nigeria. Journal of Water, Sanitation and Hygiene for Development. 2022;12(7):517–28.

99. Sommer M, Kjellén M, Pensulo C. Girls’ and women’s unmet needs for menstrual hygiene management (MHM): the interactions between MHM and sanitation systems in low- income countries. Journal of Water, Sanitation and Hygiene for Development. 2013;3(3):283–97.

100. Winter SC, Sommer M, Obara LM, Nair D. “There is no place to dispose them. What would you have me do?’’: A qualitative study of menstruation in the unique physical and social environment in informal settlements in Nairobi, Kenya. Health & Place. 2022;78:102932.

101. Elledge MF, Muralidharan A, Parker A, Ravndal KT, Siddiqui M, Toolaram AP, et al. Menstrual Hygiene Management and Waste Disposal in Low and Middle Income Countries—A Review of the Literature. International Journal of Environmental Research and Public Health. 2018;15(11):2562.

102. Wilbur J, Poilapa R, Morrison C. Menstrual Health Experiences of People with Intellectual Disabilities and Their Caregivers during Vanuatu’s Humanitarian Responses: A Qualitative Study. International Journal of Environmental Research and Public Health [Internet]. 2022; 19(21).

103. Bhattacharjee M. Menstrual Hygiene Management During Emergencies: A Study of Challenges Faced by Women and Adolescent Girls Living in Flood-prone Districts in Assam. Indian Journal of Gender Studies. 2019;26(1-2):96–107.

104. Sadique S, Ali I, Ali S. Managing menstruation during natural disasters: menstruation hygiene management during “super floods” in Sindh province of Pakistan. Journal of Biosocial Science. 2024;56(3):480–92.

105. Torondel B, Sinha S, Mohanty JR, Swain T, Sahoo P, Panda B, et al. Association between unhygienic menstrual management practices and prevalence of lower reproductive tract infections: a hospital-based cross-sectional study in Odisha, India. BMC Infectious Diseases. 2018;18(1):473.

