## Supplementary material for "The Effects of Climate Hazards on Personal Hygiene Practices among People with Disabilities in Bangladesh: A Qualitative Study": Guideline to interview people with disabilities

**Guidelines to interview people with disabilities**

**20 August 2023**

**Interview guide**

**Title of Project: Inclusive pathways towards climate-resilient WASH in Bangladesh**

*Introduce yourself, go through the information sheet, and seek and confirm consent or assent.*

*Throughout the questions, probe deeply into the issues. Explore why choices were made and reasons for changes in behaviours, how these impacted the person and the person who supports them (if relevant).*

### Washington Group short set on Functioning – Enhanced (WG-SS Enhanced^[1]^)

*Only participants who answer ‘a lot of difficulty’ or ‘cannot do at all’ across one or more functional domain can be included in this study. We are not asking the WG-SS Enhanced questions about anxiety and depression.*

*Interviewer read:* "The next questions ask about difficulties you may have doing certain activities.”

**VISION**

[Do/Does] [you/he/she] have difficulty seeing, even when wearing [your/his/her] glasses]? Would you say... [*Read response categories*]

1. No difficulty
2. Some difficulty

3. A lot of difficulty

4. Cannot do at all

*7. Refused
9. Don’t know*

**HEARING**

[Do/Does] [you/he/she] have difficulty hearing, even when using a hearing aid(s)]? Would you say... [*Read response categories*]

1. No difficulty
2. Some difficulty

3. A lot of difficulty

4. Cannot do at all

*7. Refused
9. Don’t know*

**MOBILITY**

[Do/Does] [you/he/she] have difficulty walking or climbing steps? Would you say... [*Read response categories*]

1. No difficulty
2. Some difficulty

3. A lot of difficulty

4. Cannot do at all

*7. Refused
9. Don’t know*

**COMMUNICATION**

Using [your/his/her] usual language, [do/does] [you/he/she] have difficulty communicating, for example understanding or being understood? Would you say... [*Read response categories*]

1. No difficulty
2. Some difficulty

3. A lot of difficulty

4. Cannot do at all

*7. Refused
9. Don’t know*

**COGNITION (REMEMBERING)**

[Do/does] [you/he/she] have difficulty remembering or concentrating? Would you say... [*Read response categories*]

1. No difficulty
2. Some difficulty

3. A lot of difficulty

4. Cannot do at all

*7. Refused
9. Don’t know*

**SELF-CARE**

[Do/does] [you/he/she] have difficulty with self care, such as washing all over or dressing? Would you say... [*Read response categories*]

1. No difficulty
2. Some difficulty

3. A lot of difficulty

4. Cannot do at all

*7. Refused
9. Don’t know*

**UPPER BODY**

[Do/Does] [you/he/she] have difficulty raising a 2 liter bottle of water or soda from waist to eye level? Would you say... [*Read response categories*]

1. No difficulty
2. Some difficulty
3. A lot of difficulty
4. Cannot do at all

*Refused
Don’t know*

[Do/Does] [you/he/she] have difficulty using [your/his/her] hands and fingers, such as picking up small objects, for example, a button or pencil, or opening or closing containers or bottles? Would you say... [*Read response categories*]

1. No difficulty
2. Some difficulty
3. A lot of difficulty
4. Cannot do at all

*Refused
Don’t know*

### Interview guide

Throughout the questions, probe deeply into the issues. Explore why choices were made and reasons for changes in behaviours, how these impacted the person and the person who supports them (if relevant).

### SECTION 1. Understanding disability

- Can you tell me about the challenges you face (your disability)?
- Do you get any support (healthcare, assistive device, WASH) from any organisation? Probe into:
  - Do you have a disability ID card? What is your ‘disability group’?
  - What kind of support do you get?
  - Are you a member of an OPD?
  - Are you a member of a WASH committee?
- Does someone support you with day-to-day activities?
  - Who?
  - What kind of support do they give you [explore WASH-related tasks]?
  - Do you usually get as much support as you need?
- Is your caregiver always around to help you collect drinking water, go to the toilet, wash your hands and body?
  - What do you do if they are not around to help?

### SECTION 2. Climate hazards

***Purpose: Participants focus on climate risks to water, sanitation and hygiene. These include fast-onset climate events (e.g. cyclone, heatwaves, flash flooding) and slow onset (e.g. sea level rise, salinisation, rainfall uncertainty and drought, increased rainfall). It does not include weather that is normal or expected, such as localised flooding after monsoon rains. Please encourage participants to focus on fast or slow-onset climate events and how these impact their access to water points, latrines and bathing facilities for personal hygiene.***

- In the last one to five years, have you experienced extreme weather events like extremely wet/rainy weather, extremely hot and dry weather, cyclones/storms, storm surge or very high tide (which are getting worse due to sea level rise)? Probe into:
  - The range of weather events experienced.
  - The extent that these events affected them.
- How did these affect you and your family? Probe into:
  - Disruption to livelihoods, schooling.
  - Access to food and shelter.
  - Safety/security.
  - Water
  - Going to the toilet
  - Handwashing facility

Bathing facility

***Now refer to these climate risks to WASH throughout the interview. Remember to steer the participant away from expected or normal weather events.***

**SECTION** **2.1 Water**

- What type of water source do you use? Probe into technology, location, distance from home.
- Did anything happen during or after the event that made your ability to reach the water source more difficult? Probe into:
  - Personal injury or illness.
  - The caregiver sustained an injury, illness, died.
  - Increased difficulties in leaving the home.
  - Damaged path or route to the water point.
  - Any other difficulties faced getting to the water point, e.g. extreme heat, dusty, slippery, muddy, flooding.
- Did anything happen to your water source (e.g. damaged, deterioration of water quality)? If yes, what happened to your water source?
- How did you adapt or manage? Probe into:
  - What alternative did you use?
  - Did you or someone else decide how you adapted and managed?
- Did you use an alterative water source? Probe into: technology, location, distance from home.
- Did you face any challenges when using the alternative water source? Probe into:
  - Getting to the water point (distance, terrain).
  - Ability to collect water independently?
  - If they had to pay a water fee.
  - Were they allowed to use the water point, or did they need permission? And if that caused problems for them.
- Did you change any of your daily routines or practices because of the disruption to your water source? Probe into:
  - Used less water for drinking, bathing, handwashing, laundry
  - Drank less, bathed less
  - The impacts of these changes on their ability to cope, work, and contribute to the family.
  - Ability to leave the home, socialise with / eat with others outside the home.
  - Ability to go to the religious institution, weddings, festivals, celebrations etc.
  - How did this affect you (make you feel)?
- Did you or anyone else repair the damaged water point? Did it help? Probe into:
  - Who repaired it?
  - Did you have a say in how that was repaired?
  - Usability, accessibility, affordability?
- If you didn’t repair it, what happened? How did you manage?
- Can you think of anything that could be done to improve your access to water all year round, including during extreme weather events?

**SECTION** **2.2 Sanitation**

- What type of latrine do you use? Probe into technology, location, distance from home.
- Did anything happen during or after the event that made your ability to reach the toilet [or place you go to the toilet] more difficult? Probe into:
  - Personal injury or illness.
  - Caregiver sustained an injury or illness.
  - Became incontinent / incontinence worsened.
  - Increased difficulties in leaving the home.
  - Damaged path or route.
  - Any other difficulties faced getting to the toilet, e.g. extreme heat, dusty, slippery, muddy, flooding.
- Did the weather event damage your toilet facility? How? Probe into:
  - Type of toilet.
  - Damage to the superstructure and latrine infrastructure/sanplat.
  - Toilet becomes backed up / stops flushing (this happens if the pit/tank is full of water)
  - Water goes over the toilet.
  - Pit overflow.
- Where did you go to the toilet after the damage? Probe into:
  - What ‘toilet’ was used – e.g. open defecation, defecating inside the home (e.g. flying toilet, bedpan, commode), used pads/diapers, neighbours toilet
  - Did you decide what toilet to use, or was it someone else? Who?
- How easy or difficult was it to use that toilet? Probe into:
  - Ease/difficulty in independent toileting.
  - Contact with urine and faeces.
  - Safety, security, privacy, dignity.
  - If they had to pay to use the toilet.
  - If they needed permission to use the toilet and if that caused any problems for them.
  - Differences between where the participant went to the toilet and non-disabled family members and why.
- How did the damage to your toilet/difficulties reaching the toilet affect your daily toileting routines or practices? Probe into:
  - Limiting toileting/holding off going to the toilet
  - Ability to go as often as required.
  - Restriction of food /water consumption with reasons why
  - Reliance on a caregiver
  - Reduced ability to leave the home.
  - Emptied the pit into the floodwater.
  - The impacts of these changes on their ability to cope, work, and contribute to the family.
  - Creates anxiety / stress / worry.
- How did this impact your health, comfort and ability to interact with others? Probe into:
  - Diarrhoea, typhoid, dysentery, cholera?
  - Ability to leave the home, socialise with / eat with others outside the home.
  - Ability to go to the religious institution, weddings, festivals, celebrations etc.
  - How did this affect you (make you feel)?
- Did you or anyone else repair the damaged toilet? Did it help? Probe into:
  - Who repaired it?
  - Usability, accessibility, safety, security, affordability
  - Did you have a say in how it was repaired?
- If you didn’t repair it, what happened? How did you manage?
- Can you think of anything that could be done to improve your access to your latrine all year round, including during extreme weather events?

**SECTION** **2.3 Personal hygiene (handwashing, anal cleansing, menstrual hygiene, bathing, laundry)**

- What water do you use for bathing? Probe into:
  - Do you use the same water for bathing as you do for drinking?
  - Is the water you use for bathing salty?
- What kind of bathing facility do you use? Probe into:
  - Type of bathing facility and location
  - If water is inside the facility
  - If the person relies on another person to bring water to the bathing facility
  - If the facility supports the participant’s privacy and dignity
  - Accessibility and useability
- How did the weather event affect your ability to maintain personal hygiene (e.g. handwashing, anal cleansing, menstrual hygiene, bathing, laundry)? Probe into:
  - Usability, privacy, safety, security
  - Damage to the path/route to the handwashing station, bathing facility, laundry area meaning [name] can no longer reach the facility.
  - Damage to the handwashing station, bathing facility, laundry area meaning [name] can no longer use them independently.
- If your ability to maintain personal hygiene was disrupted, how did it affect your health, comfort and ability to interact with others? Probe into:
  - Increased skin complaints, pressure sores/lesions?
  - Ability to leave the home, socialise with / eat with others outside the home.
  - Ability to go to the religious institution, weddings, festivals, celebrations etc.
  - How did this affect you (make you feel)?
- Did you or anyone else repair the handwashing station or bathing shelter? Did it help? Probe into:
  - Who repaired it and why?
  - Usability, accessibility, safety, security
  - Did you have a say in how it was repaired?
- Did you or anyone else repair the damaged water point? Did it help? Probe into:
  - Who repaired it?
  - Usability, accessibility, affordability
- If you didn’t repair it, what happened? How did you manage?
- Can you think of anything that could be done to ensure you can maintain personal hygiene all year round, including during extreme weather events?
- Is it ok to ask you some questions about menstruation?
- Do you menstruate?
- What menstrual materials do you use generally? Probe into:
  - If a combination of materials are used (e.g. cloth and commercial pad) and reasons for this?
- Do you use the same menstrual material during a weather event? Probe into:
  - Reasons for using different materials? (E.g. unable to dry cloths quickly enough in heavy rains, received specific materials hygiene packs distributed in an emergency)
  - Satisfaction with materials used during a weather event? (E.g. levels of comfort, confidence in the material’s absorbency etc)
- Did the weather event affect your ability to manage menstruation in other ways? How and what did you do in response? Probe into:
  - Ability to wash/dry and dispose of the menstrual material?
  - Frequency of changing the menstrual material
  - Ability to bathe and wash thoroughly with clean water?
  - Any skin complaints, e.g., itching, sore vagina, or surrounding skin?
- Can you think of anything that could be done to improve your ability to manage menstruation all year round, including during extreme weather events? Probing question:
  - Having an emergency hygiene kit/pack that includes various types of menstrual materials.

### SECTION 3. Access to disaster preparedness information

***Purpose: to understand if participants a) get weather forecasts and disaster warnings and b) if access to that information has helped them prepare for the event. If they do not get that information, how has it impacted their ability to prepare?***

- Do you have access to weather forecasts? If yes, probe into:
  - Source of this information? (E.g. friends, family, radio announcements, TV, posters, loudspeaker, community volunteer, OPD)
  - If it is understandable/accessible?
  - If they do anything differently because of the weather forecasts?
- If no, probe into:
  - If other people get weather forecasts?
  - Why don’t they get this information?
  - How this lack of information affects them?
  - What could be done to improve this?
- Do you have access to disaster warnings? If yes, probe into:
  - Which types of disasters do you get warnings about (e.g. cyclone, flood, drought)?
  - Source of this information? (E.g. friends, family, radio announcements, TV, posters, loudspeaker, community volunteer, OPD)
  - If it was understandable/accessible?
  - Do you have information about where to go in a disaster (e.g. shelter)?
  - What did they do after the warning?
  - Did they need assistance to act after the warning?
  - Were they given assistance? Who helped them?
- If no, probe into:
  - Are there warnings at the Upzilas level but not at the union/village level?
  - If other people get warnings?
  - Why don’t they get this information?
  - How this lack of information affects them?
- What kind of weather warnings would be helpful? Probe into:
  - When it would be useful to get them?
  - How they would like to receive them?
  - What information would you find helpful and why?
- Do you or your family have a disaster plan, evacuation checklist, evacuation pack? Probe into:
  - What does this include?
  - Were you consulted when this was developed?
  - Are your needs/requirements considered? How?
- Can you think of anything you’d like support with to help you prepare for a weather event? Probe into:
  - The development of a family disaster plan.
  - Understanding what to put in an evacuation pack.
  - Having an evacuation checklist.
  - Helping family members understand how to support a person with disabilities during a weather event.
  - A list of telephone numbers for disability support services (e.g. medical support, assistive devices and products).

### SECTION 4. Assistance

***Purpose: to explore assistance provided to persons with disabilities, what that enabled them to do, if their assistive devices (e.g. wheelchair, glasses) were damaged in a weather event, and if they got them fixed or replaced; if not, how this impacted their independence.***

- Did you get assistance during/after a weather event? If yes, probe into:
  - What assistance was it (WASH, assistive device, support to reach a shelter)?
  - Who provided assistance? (E.g. the government, other organisations, village leader, friend, family)
  - How did you get it? E.g. directly, through your family, applied for it, met the criteria?
  - Did the assistance help?
- Did you have a say about how the assistance was used? Probe into:
  - Could you personally use any of the assistance provided?
  - If not, why not?
- What did you do with that assistance?
- If you or your family did not get any assistance during/after a weather event, why not? Probe into:
  - Did your neighbours get assistance? If so, why did they get assistance, and you did not?
- Is there any assistance that you didn’t get that would be helpful?
- If you use an assistive device, has it ever been damaged during a weather event? If yes, probe into:
  - What assistive device and how it was damaged?
  - What happened? How did it impact you / your caregiver?
  - Did you try to mend it or get a new device?
  - Whom did you approach for that? What happened?
  - If not, how did you manage?

### SECTION 5. Capacity

***Purpose: This section explores what priority people put on maintaining WASH services during/after a weather event, the activities the community, household and individual do to protect their WASH services from a weather event and the ability of the person with disabilities to participate in those activities.***

- Earlier, you told me about how extreme weather events affect you and your family’s ability to (work, grow/buy food, go to school etc.). You’ve also told me how the events affect your ability to access water, toilets and maintain personal hygiene. Thinking about all these issues, how do you and your family prioritise fixing or maintaining access to water, toilets, and bathing facilities compared to the other priorities?
- In your family and village, who decides how to prepare for and respond to weather events?
- Are you involved in the decision-making? If yes, how? If not, probe into:
  - Why are you not involved?
  - Do you want to be involved? How and why?
- In your village, what activities do people do to prepare for extreme weather events? Probe into:
  - Clearing drains, rivers and streams.
  - Monitoring groundwater levels and surface flow.
  - Mending/maintaining tubewells.
  - Increasing water storage, constructing rainwater harvesting.
  - Reinforcing latrines, emptying latrines more regularly before the monsoon.
  - Constructing more sustainable latrines (on / off-site septic tanks, ecosan etc).
  - Reinforcing handwashing messages and behaviours.
  - If anyone in the community shares information and/or resources with others.
- Are you ever involved in these activities? If yes, probe into:
  - Their role and the activities they carry out.
  - How they felt about doing these activities (e.g. gave them confidence, ability to interact with different people etc.).
  - Who mobilised them?
  - If they were supported to carry out the activities if required.
  - How others reacted to their involvement.
- If no, probe into:
  - Would you like to be involved?
  - Reasons for that lack of involvement.
  - Desire to be involved and why.
  - What support would be required to enable their involvement?
- Are you involved in the disaster management or shelter management committee? Probe into:
  - What do these committees do?
  - What is your role within that?
  - Have you made decisions?

### SECTION 6. Visioning

**Purpose: To encourage the participants to think about solutions. These will contribute to the participatory workshop where we co-develop recommendations.**

- Beyond what you have already told me, what could be done to help you have better access to water, toilets and hygiene during or after weather events? Probe into:
  - Ability to access water points, toilets, and bathing facilities independently.
  - Ability to manage menstruation comfortably.
  - Accessing weather forecasts and disaster preparedness information
  - Accessing assistance, including assistive devices
  - Improving the community, their family, and their own ability to prepare, respond and recover from a weather event
- Thank you very much for talking to me about your experiences. Before we finish, is there anything else you want to tell me? Is there anything you would like to ask me?

1. <https://www.washingtongroup-disability.com/question-sets/wg-short-set-on-functioning-enhanced-wg-ss-enhanced/>
