## Supplementary material for "The Effects of Climate Hazards on Personal Hygiene Practices among People with Disabilities in Bangladesh: A Qualitative Study": Guideline to interview caregivers

**21 Aug 2023**

**Interview guide: Caregivers**

**Title of Project: Inclusive pathways towards climate-resilient WASH in Bangladesh**

*Introduce yourself, go through the information sheet, and seek and confirm consent or assent.*

*Throughout the questions, probe deeply into the issues. Explore why choices were made and reasons for changes in behaviours, how these impacted them.*

### Interview guide

Throughout the questions, probe deeply into the issues. Explore why choices were made and reasons for changes in behaviours, how these impacted the person and the person who supports them (if relevant).

### SECTION 1. Understanding caregiving

***Purpose: To understand the person with disabilities’ support requirements and WASH care tasks performed.***

- What is your relationship with the person you support? Can you tell me about them? Probe into:
  - Functional limitations
  - Their main caregiving tasks
  - If someone else supports them in their caregiving role
- What WASH tasks do you support [name] with? Probe into:
  - Water for drinking, personal hygiene, laundry
  - Toileting: bedpans, taking the person to the toilet, supporting them when using the toilet
  - Personal hygiene: bathing, menstrual hygiene, hand hygiene
  - Lifting/carrying to and from the WASH facility

### SECTION 2. Climate hazards

***Purpose***: ***Participants focus on climate risks to water, sanitation and hygiene. These include fast-onset climate events (e.g. cyclone, heatwaves, flash flooding) and slow onset (e.g. sea level rise, salinisation, rainfall uncertainty and drought, increased rainfall). It does not include weather that is normal or expected, such as localised flooding after monsoon rains. Please encourage participants to focus on fast or slow-onset climate events and how these impact their access to water points, latrines and bathing facilities for personal hygiene, their ability to support the person with disabilities and their own wellbeing.***

- In the last one to five years, have you experienced any extreme weather events like extremely wet/rainy weather, extremely hot and dry weather, cyclones/storms, storm surge or very high tide (which are getting worse due to sea level rise)? Probe into:
  - The type of weather event (fast or slow onset).
  - The range of weather events experienced.
  - The extent that these events affected them.
- How did these affect you and your family? Probe into:
  - Disruption to livelihoods, schooling.
  - Access to food and shelter.
  - Safety/security.
- Did any of these extreme weather events affect your ability to collect water, go to the toilet, wash your hands and bathe?

***Now refer to these climate risks to WASH throughout the interview. Remember to steer the participant away from expected or normal weather events.***

**SECTION** **2.1 Water**

- What type of water source do you use? Where is it?
- Did anything happen during or after the event that made your ability to reach the water source more difficult? Probe into:
  - Personal injury or illness
  - Person with disabilities sustained an injury or illness
  - Damaged path or route to the water point
  - Any other difficulties faced getting to the water point, e.g. extreme heat, dusty, slippery, muddy, flooding.
- Did anything happen to your water source (e.g. damaged, deterioration of water quality)? If yes, what happened to your water source?
- How did you adapt or manage? Did you use an alternative? Probe into:
  - What alternative did you use?
- Did you face any challenges when using the alternative water source? Probe into:
  - Getting to the water point (distance, terrain).
  - If they had to pay a water fee.
  - Were they allowed to use the water point, or did they need permission? And if that caused problems for them.
- Did this affect how you support [name] in meeting their water use needs? Probe into:
  - Multiple trips to the water point in the heat, rain, wind
  - If these water tasks take longer than when there is no weather event
  - What this means for their ability to carry out other tasks, such as agriculture, income generation, ability to socialise outside the home
  - Bathe [name] less, do less laundry, drink less / provide less drinking water

How did this affect you (make you feel)?

- Can you think of anything that could be done to improve your access to water all year round, including during extreme weather events? Probe into:
  - How would this enable you to support [name’s] water needs

**SECTION** **2.2 Sanitation**

- What type of toilet does [name] use? Probe into:
  - [Name’s] ability to use it independently
  - Support the caregiver provides to support [name’s] toileting
- Did anything happen during or after the weather event to make your ability to support [name’s] toileting requirements more difficult? Probe into:
  - Personal injury or illness
  - [Name] sustained an injury or illness
  - [Name] became incontinent / their incontinence worsened
  - Lifting device / commode / bedpan was lost or damaged
  - Path to the toilet was damaged
  - Any other difficulties [name] faced getting to the toilet, e.g. extreme heat, dusty, slippery, muddy, flooding.
- Was the toilet [name] uses damaged? Probe into:
  - How it was damaged (e.g. damage to the superstructure and latrine infrastructure)
  - Toilet becomes backed up / stops flushing (this happens if the pit/tank is full of water)
  - Water goes over the toilet
  - Pit overflow
  - If this impacted the caregiver’s ability to support [name’s toileting requirements]
- Where did you take [name] to the toilet after the damage? Probe into:
  - What ‘toilet’ was used – e.g. open defecation, defecating inside the home (e.g. flying toilet, bedpan), used pads/diapers
  - Did [name] or someone else decide where to go to the toilet? Who?
- How do you cope and support [name’s] toileting during these times?
  - Ease/difficulty in supporting toileting, including lifting and bathing
  - Contact with urine and faeces
  - Ability to maintain the person with disabilities’ safety, security, privacy, and dignity
  - Ability to take assist the person with disabilities as often as required
  - Any differences between where the person with disabilities goes to the toilet and the rest of the family who don’t have disabilities
  - If [name] had to pay to use the toilet.
  - If they needed permission to use the toilet and if that caused any problems for them.
- What did the damage to your toilet/difficulties in [name] reaching the toilet affect your ability to:
  - Leave the home, socialise with / eat with others outside the home.
  - Ability to go to the mosque, weddings, festivals, celebrations etc.
  - Balance caring duties with work, and contribute to the family and community
  - How did this affect you (make you feel)?
- Did you or anyone else repair the damaged toilet? Did it help? Probe into:
  - Who repaired it?
  - The ability of the caregiver to support [name’s] toileting
  - Usability, accessibility, safety, security for [name] and them when providing support
- Can you think of anything that could be done to improve your access to your latrine all year round, including during extreme weather events?

**SECTION** **2.3 Personal hygiene (supporting name’s handwashing, anal cleansing, menstrual hygiene, bathing, laundry)**

- What kind of bathing facility does [name] use? Probe into:
  - Type of bathing facility and location
  - If water is inside the facility
  - If [name] relies on the caregiver to take water to the facility
  - If it supported the privacy and dignity of the person with disabilities
  - Ability of the caregiver to use it with the person with disabilities
- How did the weather event affect your ability to support [name’s] personal hygiene? Probe into: Probe into:
  - Usability, privacy, safety, security
  - Damage to the path / route to the handwashing station, bathing facility, laundry area meaning [name] can no longer reach the facility
  - Damage to the handwashing station, bathing facility, laundry area meaning [name] can no longer use them independently
- How did the damage to your handwashing station or bathing facility affect your ability to support [name’s] personal hygiene? Probe into:
  - Washed [name’s] hands less
  - Bathed [[name] less
  - Did [name’s] laundry less
  - Need to provide more support to [name]
  - How these changes affected the caregiver’s ability to cope, work, and contribute to the family
  - How did this affect you/make you feel?
- Did you or anyone else repair the handwashing station or bathing shelter? Did it help? Probe into:
  - Who repaired it
  - The ability of the caregiver to support [name’s] personal hygiene
  - Usability, accessibility, safety, and security for [name] and them when providing support
- Can you think of anything that could be done to ensure you can maintain [name’s] personal hygiene all year round, including during extreme weather events?
- Does [name] menstruate? Do you support [name] when they are menstruating? ***[If yes, continue]***
- Did the weather event change how you supported [name] when menstruating? Probe into:
  - Accessing menstrual materials for [name]
  - Ability to wash/dry and dispose of the menstrual material?
  - Frequency of changing the menstrual material
  - Ability to bathe and wash [name] thoroughly with clean water?
  - Has [name] had any skin complaints, e.g., itching, sore vagina, or surrounding skin?
- Can you think of anything that could be done to improve your ability to support [name] to manage menstruation all year round, including during extreme weather events?

### SECTION 3. Access to disaster preparedness information

***Purpose: to understand if participants a) get weather forecasts and disaster warnings and b) if access to that information has helped them prepare for the event. If they do not get that information, how has it impacted their ability to prepare?***

- Do you have access to weather forecasts? If yes, probe into:
  - Source of this information? (E.g. friends, family, radio announcements, TV, posters, loudspeaker)
  - If it is understandable/accessible?
  - If they do anything differently because of the weather forecasts?
  - If and how they support [name] to prepare?
- If no, probe into:
  - If other people get weather forecasts?
  - Why don’t they get this information?
  - How this lack of information affects them?
  - How this affects their ability to support [name] to prepare and how it impacts on [name]?
  - What could be done to improve this?
- Do you have access to disaster warnings? If yes, probe into:
  - Which types of disasters do you get warnings about (e.g. cyclone, flood, drought)?
  - Source of this information? (E.g. friends, family, radio announcements, TV, posters, loudspeaker)
  - If it was understandable/accessible?
  - What did they do after the warning?
  - If / how they support [name] to prepare for the disaster?
  - Did they need assistance to act and support [name] to act after the warning?
  - Were they given assistance? Who helped them?
- If no, probe into:
  - Are there warnings at the Upzilas level but not at the union/village level?
  - If other people get warnings?
  - Why don’t they get this information?
  - How this lack of information affects them and [name]?
- What kind of weather warnings would be helpful? Probe into:
  - When it would be useful to get them?
  - How they would like to receive them?
  - What information would you find helpful and why?
- Can you think of anything you’d like support with to help you ane [name] prepare for a weather event? Probe into:
  - The development of a family disaster plan that includes a person with disabilities.
  - Understanding what to put in an evacuation pack.
  - Having an evacuation checklist.
  - Helping family members understand how to support a person with disabilities during a weather event.
  - A list of telephone numbers for disability support services (e.g. medical support, assistive devices – e.g. wheelchair, glasses, and products – e.g. commodes, bedpans, lifting devices).

### SECTION 4. Assistance

***Purpose: to explore assistance provided to persons with disabilities and their families/caregivers, what that enabled the caregiver to do, if the person with disabilities’ assistive devices (e.g. wheelchair, glasses) were damaged in a weather event, and if they got them fixed or replaced; if not, how this impacted their independence and the caregiver’s ability to provide support. If the caregiver uses an assistive product (e.g. lifting belt, commode), if this was damaged in the weather event and the impact of that.***

- Did you or [name] get assistance during/after a weather event? Probe into:
  - Who provided assistance? (E.g. the government, other organisations, village leader, friend, family)
  - What assistance was it?
  - How did you get it? E.g. directly, through your family, applied for it, met the criteria?
- What did you do with that assistance? Probe into:
  - How this helped the caregiver support [name].

Is there any assistance that you didn’t get that would be helpful?

- Do you use assistive products (e.g. commode, lifting belt, bathing chair etc)? If yes, was it damaged during a weather event? Probe into:
  - What happened? How did it impact your ability to support [name]?
  - If the caregiver was more likely to come into contact with urine and faeces?
  - How it impacted their physical health (e.g. back pain, water-related diseases)?
  - Did you try to mend it or get a new product?
  - Whom did you approach for that? What happened?

### SECTION 5. Capacity

***Purpose: This section explores what priority people put on maintaining WASH services during/after a weather event, the activities the community, household and individual do to protect their WASH services from a weather event and the ability of the person with disabilities and caregiver to participate in those activities.***

- Earlier, you told me about how extreme weather events affect you and your family’s ability to (work, grow/buy food, go to school etc.). You’ve also told me how the events affect your ability to support [name] to access water, toilets and maintain personal hygiene. Thinking about all these issues, how do you prioritise fixing or maintaining access to water, toilets, and bathing facilities compared to the other priorities?
- In your family and village, who decides how to prepare for and respond to weather events?
- Are you involved in the decision-making? If yes, how? If not, probe into:
  - Why are you not involved?
  - Do you want to be involved? How and why?
- In your village, what activities do people do to prepare for extreme weather events? Probe into:
  - Clearing drains, rivers and streams
  - Monitoring groundwater levels and surface flow
  - Mending/maintaining tubewells
  - Increasing water storage, constructing rainwater harvesting
  - Reinforcing latrines, emptying latrines more regularly before the monsoon
  - Constructing more sustainable latrines (on / off-site septic tanks, ecosan etc)
  - Reinforcing handwashing messages and behaviours
  - If anyone in the community shares information and/or resources with others.
- Are you ever involved in these activities? If yes, probe into:
  - If [name] came with them or if [name] was supported by someone else?
  - Their role and the activities they carry out
  - How they felt about doing these activities (e.g. gave them confidence, ability to interact with different people etc)
  - Who mobilised them?
- If no, probe into:
  - Would you like to be involved?
  - Reasons for that lack of involvement
  - Desire to be involved and why
  - What support would be required to enable their involvement?
- Can you think of any activities that could be done by you, your family, your community to prepare, respond or recover from a weather event? Probe into:
  - How they think [name] could contribute to that

### SECTION 6. Visioning

**Purpose: To encourage the participants to think about solutions. These will contribute to the participatory workshop where we co-develop recommendations.**

- Beyond what you have already told me, what could be done to help [name] have better access to water, toilets and hygiene during or after weather events and your ability to support them? Probe into:
  - Ability to access water points, toilets, and bathing facilities independently.
  - Ability to manage menstruation comfortably.
  - Accessing weather forecasts and disaster preparedness information
  - Accessing assistance, including for assistive products
  - Improving the community, their family, and their own ability to prepare, respond and recover from a weather event
- Thank you very much for talking to me about your experiences. Before we finish, is there anything else you want to tell me? Is there anything you would like to ask me?
