## Supplementary material for "The Effects of Climate Hazards on Personal Hygiene Practices among People with Disabilities in Bangladesh: A Qualitative Study": Guideline for accessibility and safety audit with people with disabilities

This draws on several data collection tools, including WaterAid (1), Wilbur et al (2, 3), and Kohlitz et al (4).

**Overview**

Participants can be people with disabilities or caregivers of people with disabilities. Complete this activity with one participant and their caregiver (if applicable) or a family member. One facilitator will lead the walk, another will be the note-taker. Do not take longer than 30 hour, including time for the person with disabilities to take breaks. Repeat the checklist for the water point, latrine, bathing shelter and handwashing station or as much as you can do in 30 minutes. Spend 30 minutes, including breaks, discussing solutions. You might not be able to complete every section with every participant. Do as much as you can but do not go over time, unless the person explicitly agrees to give you more time.

**Roles**

*Facilitator*:

1. Walk with the participants to the facility.
2. During the walk, refer to Checklist 1.
3. Once you reach the facility, refer to Checklist 2. Say what you are doing (e.g. walking to the water point) and observe (e.g. watching the participant get into the toilet, any trip hazards etc) for the voice recorder.
4. Capture answers the participant gives you on the voice recorder. Ensure the voice recorder captures these clearly.

*Note-taker*:

- Take additional notes about things the voice recorder cannot capture.
- Check that the facilitator has covered every part of the checklist.

**Materials needed**

- Checklists 1 & 2: Notebook, pen, voice recorder
- Satisfaction scale and discussing solutions: Large piece of paper, marker pen

**Climate risks to WASH only**

Participants must focus on climate risks to water, sanitation and hygiene. These include fast-onset climate events (e.g. cyclone, heatwaves, flash flooding) and slow onset (e.g. sea level rise, salinisation, rainfall uncertainty and drought, increased rainfall). It does not include weather that is normal or expected, such as localised flooding after monsoon rains. Please encourage participants to focus on fast or slow-onset climate events and how these impact their access to water points, latrines and bathing facilities for personal hygiene.

Remember to repeat the checklist for the water point, latrine, bathing shelter and handwashing station or as much as you can do in 30 minutes. Spend 30 minutes, including breaks, discussing solutions.

**Explain the purpose of the activity to the participant**

The purpose of this activity is to understand the access barriers to water, sanitation and personal hygiene facilities faced by people with disabilities and how climates, like very hot and dry weather, heavy rainfall, or cyclones, also affect these barriers. We will go to your toilet, water point, bathing facility and handwashing station so you can show me how you get there and use them. I’m very interested to hear about any solutions you think would help. We will spend time after we’ve seen the facilities talking about these solutions.

**SECTION 1. CLIMATE HAZARDS**

***Purpose: Participants focus on climate risks to water, sanitation and hygiene. These include fast-onset climate events (e.g. cyclone, heatwaves, flash flooding) and slow onset (e.g. sea level rise, salinisation, rainfall uncertainty and drought, increased rainfall). It does not include weather that is normal or expected, such as localised flooding after monsoon rains. Please encourage participants to focus on fast or slow-onset climate events and how these impact their access to water points, latrines and bathing facilities for personal hygiene.***

- In the last one to five years, have you experienced extreme weather events like extremely wet/rainy weather, extremely hot and dry weather, cyclones/storms, storm surge or very high tide (which are getting worse due to sea level rise)? Probe into:
  - The range of weather events experienced.
  - The extent that these events affected them.
- How did these affect you and your family? Probe into:
  - Disruption to livelihoods, schooling.
  - Access to food and shelter.
  - Safety/security.
- Did any of these extreme weather events affect your ability to collect water, go to the toilet, wash your hands and bathe?

***Now refer to these climate risks to WASH throughout the interview. Remember to steer the participant away from expected or normal weather events.***

**SECTION 2. CHECKLISTS**

**Checklist 1 – The journey to the facility**

| **Question** | **How do weather events affect this** **(e.g. very hot and dry weather, heavy rainfall, floods, or cyclones)? Encourage the participant to consider the last climate disruption experienced. Prompts include:** | **Solutions** |
| --- | --- | --- |
| What type of water point / latrine / bathing facility are we going to see? Has anything been done to it better able to withstand a weather event? Please explain. |  |  |
| Is there anything blocking the path that could make you trip or makes it more difficult to use? | Is there increased debris after a flood/heavy rain? |  |
| Is the path wide enough for you (and your caregiver) to use? |  |  |
| Are there any parts of the path that are steep or slippery? |  |  |
| During the night, is it difficult to get there? |  |  |
| Are there any times you feel unsafe getting to the facility? When/why? |  |  |
| Is the distance to the facility far for you? | Is it difficult to get to the facility in a very hot day, or when it rains heavily?  Is it difficult to carry water when it is very hot or during heavy rains? |  |
| If the facility is not working, how far is the next facility and how difficult is it to get to? | Do heavy rains or dry periods cause the facility to stop working? |  |
| Do you use something to help you transport water back home (e.g. trolley, rack under the seat of a tricycle or wheelchair, hook attached to the crossbar of a crutch)? | Do heavy rains or dry periods make it more difficult to transport water? |  |

**Checklist 2**

A. **Accessibility of the sanitation facility**

| **Question** | **How does the climate affect this? (e.g. floods / water clogging because of heavy rain, after a storm, drought, or a cyclone)** | **Solutions** |
| --- | --- | --- |
| How easy or difficult is it to enter the facility? Is the entrance wide enough? |  |  |
| Is there enough space inside for you (and your caregiver) to move around easily? |  |  |
| Is the floor smooth and dry? | When there is heavy rain, does the ground get very wet or muddy? |  |
| Are there any handles /handrails/ structures to prevent you from falling? |  |  |
| Is the facility structure robust and private? | Under windy conditions, will the walls or roof blow away? |  |
| Does the facility have a door and lock that you can use independently? | Under windy conditions, will the door stay in place? |  |
| Is it light enough inside? |  |  |
| Are there any times you feel unsafe using the facility (e.g. at night)? When/why? |  |  |
| Is the facility uncomfortable to use? Why? | Does the toilet smell worse when it is very hot or there is heavy rain?  When it floods, does the water cover the latrine? |  |
| Can you reach the water for anal cleansing? Who collects this water? | Is there always water available for personal hygiene during dry spells? |  |
| Are the areas inside and around the facility clean and acceptable quality? | Does this change during heavy rains or cyclones? |  |
| Is the handwashing station and soap/ash easy for you to use? | Is there always water available even during / after heavy rains, dry spells, cyclones? |  |
| **MENSTRUAL HEALTH** | | |
| What menstrual material/s do you use? |  |  |
| In the weather event, did you use a different menstrual material? Why? |  |  |
| Where do you change your menstrual material? |  |  |
| Is there water and a private space to wash your body when you are menstruating? | Is there always water available even during / after a climatic event? |  |
| Is there space to change a menstrual material, wash and dry a menstrual material? Is that difficult for you? | How can you dry it in heavy rains? What do you do? |  |
| If you dispose of a menstrual material, where do you put it? Is that difficult for you? |  |  |

B. **Accessibility of the water point**

| **Question** | **How does the climate affect this? (e.g. floods / water clogging because of heavy rain, after a storm, drought, or a cyclone)** | **Solutions** |
| --- | --- | --- |
| Is the floor surrounding the water point smooth and dry? | When there is heavy rain, does the ground get very wet or muddy? |  |
| Are there any handles /handrails/ structures to prevent you from falling? |  |  |
| Is the water point difficult to use? Why? | Do these difficulties increase during floods or droughts? |  |
| Is water always available? | How is water quantity and quality affected by an extreme weather event? |  |
| Is there a structure to help you lift water containers from the floor (e.g. pedestal at knee  water container resting stand/table so you don’t need to bend so low to pick up the water container)? | Can you still use these extreme weather events? |  |
| Are there any times you feel unsafe using the facility? When/why? |  |  |

**SECTION 3. SATISFACTION**

**Satisfaction scale**

After completing the checklists for one facility, complete the satisfaction scale. Repeat this for every facility audited.

How satisfied are you with this facility when there is a weather event (e.g. sea level rise, salinisation, rainfall uncertainty and drought, increased rainfall)?

| Unsatisfied | Somewhat satisfied | Satisfied |
| --- | --- | --- |
| 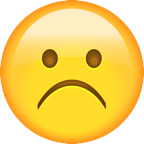 | 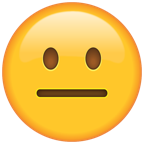 | 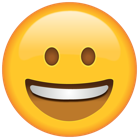 |

***Ask the participant to select one of the three options. Say which answer the participant selected into the voice recorder.***

How satisfied are you with this facility when there is a disaster (e.g. flood, cyclone)?

| Unsatisfied | Somewhat satisfied | Satisfied |
| --- | --- | --- |
| 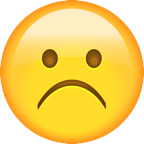 | 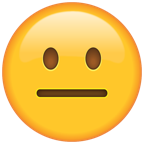 | 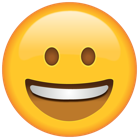 |

***Ask the participant to select one of the three options. Say which answer the participant selected into the voice recorder.***

**SECTION 4. SOLUTIONS**

**Discussing solutions**

This must be completed with people who completed the Accessibility and Safety Audit. Explain that you will move to identifying and developing solutions to the problems highlighted in the audit (checklists 1 and 2).

1. Draw Table 1 on a large piece of paper.
2. Ask the participants about the issues in checklists 1 and 2. Write these down in Table 1. Draw pictures to depict the issues.
3. Ask the participants about the solutions discussed in checklists 1 and 2. Write these down in Table 1.
4. Ask the participants responsible for implementing each solution: household/family, community, or local government.
5. Write down each solution the participant mentions under the column heading of who is responsible for implementing it. Some solutions can be written under two or all three headings if needed.

***Nb. Remind participants to think about problems and solutions relating to accessibility during fast-onset climate events (e.g. cyclone, heatwaves, flash flooding) and slow onset events (e.g. sea level rise, salinisation, rainfall uncertainty and drought, increased rainfall).***

Table 1. Solution planning

| Issue  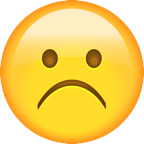 | Solution  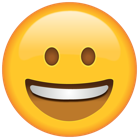 | Household/family 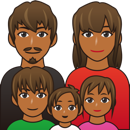 | Community  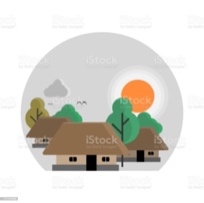 | Local government  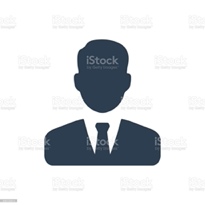 |
| --- | --- | --- | --- | --- |

**References**

1.	WEDC, WaterAid. Accessibility and safety audit: latrine. 2013.

2.	Wilbur J, Kayastha S, Mahon T, Torondel B, Hameed S, Sigdel A, et al. Qualitative study exploring the barriers to menstrual hygiene management faced by adolescents and young people with a disability, and their carers in the Kavrepalanchok district, Nepal. BMC Public Health. 2021;21(1):476.

3.	Wilbur J, Morrison C, Iakavai J, Shem J, Poilapa R, Bambery L, et al. “The weather is not good”: exploring the menstrual health experiences of menstruators with and without disabilities in Vanuatu. The Lancet Regional Health – Western Pacific. 2021.

4.	Kohlitz J, Megaw T, Chong J, Sugi F, Palaipeni P, Emanual Y, et al. Climate Change Response for Inclusive WASH: A guidance note for Plan International Indonesia. Prepared by ISF-UTS for Plan International Indonesia. Available at <https://waterforwomen.uts.edu.au/outputs/climate-outputs/ccriw-guidancenote-indonesia/> (accessed 27 March 2023); 2020.
