## Supplementary material for "The Effects of Climate Hazards on Personal Hygiene Practices among People with Disabilities in Bangladesh: A Qualitative Study": Photovoice and ranking guideline

**S5: PhotoVoice and Ranking Guideline**

### Purpose of PhotoVoice

This exercise is designed to explore 1) how climate disruptions (e.g. heavy rainfall, flooding, drought, cyclone) impact the WASH experiences of women and men with disabilities, 2) how this population attempts to manage these events, 3) how this impact their mental health and wellbeing and 4) how participants prioritise their importance.

This method aims to enable participants to take photos of their personal perspectives and experiences of how these challenges affect the lives of individuals with disabilities. This method is particularly important for exploring topics that can be difficult to talk about, and the photos bring new insights and perspectives which raise awareness about more invisible topics. Images can be used in exhibitions, online, research reports and other media channels and have been used to influence policy and decision-makers and implementers^[1]^. The PhotoVoice process used in this research was partly taken from the process developed by photovoice.org (<http://www.photovoice.org/methodologyseries/inclusivemethodology/designing.htm>).

### Requirements

- Either 1) a digital camera, SD card and spare battery, or 2) a camera phone that is fully charged
- Photo printing facilities
- Voice recorder
- Notebook and pen

### Identifying participants

It may not be possible to do PhotoVoice with all individuals with disabilities. Participants need to be able to follow instructions and think critically about the task being given. Because of the nature of digital cameras, motor skills are important. For individuals with severe intellectual disabilities, this task might be difficult. However, it has been successfully used with young people with mild-moderate intellectual impairments in Vanuatu (Wilbur et al, in draft). PhotoVoice works well with those who have physical and sensory disabilities (including those who are totally blind). Some participants may need the support of someone they trust to guide them. For instance, a person with a visual impairment may need to direct another person to arrange selected items within the frame of the photo. This activity will be done with individuals, not groups.

### Information and consent

The consent process must be done very thoroughly to ensure that the participant understands the purpose of the activity and what they are agreeing to. Two consent processes will be carried out for participants doing PhotoVoice. An initial informed consent at the start of the process, and a second informed consent after the photos have been taken and before the interview is carried out. The secondary consent relates to how the photos can be used and how they should be acknowledged. This is done after the photos are taken so that the participant can make a better judgement about how they wish them to be used.

#### First meeting

This first meeting will take up to one hour.

1. **Seek initial informed consent. If granted, continue**
2. **Understanding the camera** - A photographic camera (digital), or a camera phone will be provided to the participant. The researcher should explain simply how the device works and allow the participant to observe (or feel if they are visually impaired) where the lens is and how the shutter works (a visually impaired person can listen to the shutter noise and feel the lens while closed, so not to leave marks on the lens). The researcher should take time to teach the participant the basic features of the device, including how to switch the camera off and on, how to take a photo and how to view the photo, how to change the battery. Allow the participant to take a photo to see what it feels like. It is important that while the participant is learning about the camera, they are also holding it to begin to be familiar with how it feels. This is particularly important for people with sensory impairments so that they begin to learn how the buttons feel and how they are positioned. A useful starting point is to remind the individual always to use the wrist strap/head strap.
3. **Understanding photography** - Since many of your participants may never have seen a camera before and may have seen relatively few photos in their life, it is also important to explain the purpose of photography. A simple way to explain this is that photography can serve several purposes. You can use photography to capture a moment you want to remember as if it was real again. Give an example of this by getting the participant to take a photo of the researchers or their house, view the photo (on the device) and point out that it looks exactly like it is in real life. Then explain that you can also use photography as if it was art by arranging things in a certain way that tells a story or creates a version of reality that can be explored or questioned. Give an example of this too – if you want to take a photo of the idea of ‘hunger’ it may be hard to show this literally. But you could use symbolism to show hunger. You might have a family seated in their living room, all looks normal, but they all have empty bowls in front of them. Explain that what we will do today is use photography to tell their story and creatively express their views.
4. **Understanding the elements of Photography** –
   a. **Taking photos without showing a face to protect the participant’s identity** – people may wish to hide their own and other’s identity, so you will need to explain how to do this. Techniques include:
      i. Taking photos with the light behind the subject so the figure will be in shadow.
      ii. Focusing on something behind or in front of the subject. This means that the subject will be in soft focus
      iii. Photographing a person's shadow
      iv. Taking a photo of someone from behind (the back of their head, their head / body etc)
      v. Not taking photos of their own house
   b. **Ensuring photos protect the participant’s dignity** – explain to the participants that they can take photos of anything related to water, sanitation and hygiene. This includes menstrual hygiene and incontinence issues. But explain that photos of menstrual or incontinence ‘accidents’ will not be used. This includes clothes, bedsheets or other materials with blood, faeces or urine stains on it
   c. **Landscape/Portrait -** Shooting can be done vertically (portrait) or horizontally (landscape). Show participants how this affects the image and explain that portraits can be better when focusing on a person and landscapes can be better when you want to capture more of the environment. For people who have visual impairments can be demonstrated using a mount board window, which can be rotated and felt by the participants. A collection of tactile objects such as toys or fruit can be a good focus for this exercise – the window can be placed by the display in each position, and the difference in what is contained in the ‘photo' felt through the window.
   d. **Framing -** Explain that when taking a photograph, it is not simply a matter of pointing towards the subject but of deciding what is included in the photo – all or some of the subject, the subject and the background, the subject and what is above it etc. Tactile objects can be a useful reference for explaining this concept to someone who is visually impaired. Show the participant how to adjust the framing by using zoom.
   e. **Foreground/background -** This must be explained verbally and using their body as a reference. For example, you can ask two participants to stand one in front of the other and then explain who is in the foreground and who is in the background and what that would mean in a photograph (i.e. who would seem more important, more prominent, larger in the frame etc). Show the participant how to change the focus on the camera.
   f. **Distance (only for people with visual impairments) -** When taking a photograph, it is very important to identify the distance to the subject to ensure it is framed as desired. This can be done by reaching with or laying out a cane, measuring it with steps, or measuring with the joints, such as hands, wrists, arms and forearms. It can be very reassuring for a photographer to know how a photo of a person will be framed if taken from a distance of one cane's length, for example.
   g. **Focus/blur -** It is important to identify the area that needs to be in focus. The photographer needs to remember that she can communicate different feelings or ideas depending on what is focused on in the photograph. Here is one way the concept can be explained in a way that makes sense to someone with no sight: When one touches a glass bottle, one identifies the material, its temperature, its dimensions and every detail that makes one recognise the object as a bottle. If this is done again with a thin cloth over the bottle, the details of the bottle won't be recognised so precisely. Nevertheless, one will still know it is a bottle since some details, like its shape and size, are still recognised. This happens when one sees an image that's blurry or out of focus; one recognises what it is but cannot make out the details.
   h. **Light -** Light plays an important role in a photograph since it produces different effects, which lead to different feelings in the observer. A person in darkness, for example, may convey an experience of feeling hidden, whereas a person in bright light may convey confidence or nothing to hide. These effects must be explained fully to blind or photographers with visual impairments who might not realise the impact of the light on their work. To explain this, try to invite the participant to think about the warmth they feel on their face if they are in the sun and use this sensation to determine where the light is coming from. Also, teach the participant how to use the flash setting for dark environments.
5. **The photographic task** - Once the participant is comfortable with all this, set them their task. Take five photos of things in their day that make them happy. Then take five photos that represent their experiences and feelings related to how climate disruptions (e.g. heavy rainfall, flooding, drought, cyclone) impact their access to and use of WASH services, how they manage this and how this affects their ability to cope. Encourage the participant to focus on one specific climate disruption they experienced. Explain that it is their choice about what they photograph. If there is anything they do not want to photograph, they do not have to. Ask the participant if anything springs to mind. Help the participant make a list of these issues so that these things can be remembered. For each one, the participant should think about how they could represent the experience or feeling. If the participant needs more guidance, work through one example with them, but make sure they lead the process. Stand back whilst the participant takes the photos. Offer guidance, but don’t lead them to take specific photos.
6. **Self-Directed portraits** - It is likely that in settings where participants are unfamiliar with cameras and photography that they will be keen to be in the photos rather than just taking them. If the participant wants to be in the picture then they still had to direct the field researcher as to how they wanted the photo to look, providing direction on whether it was to be a portrait or landscape shot, what was in the foreground or background, how much of their body should be in shot etc.
7. **Arrange a suitable time to return to the individual’s house to give them their photos and have a short discussion.**

#### Second meeting

- This second meeting will take up to one hour
- Show the participant the printed photos
- Seek secondary informed consent. If given, proceed
- It is impossible to do guiding questions for PhotoVoice as we don’t know what participants will take photos of. Have a copy of the topic guides with you. If any photos are related to any of the topics explored in the interview pack, draw on the relevant questions. If they take a photo of the toilet, draw on questions related to this in the topic guide or the accessibility and safety audit.

**INSTRUCTIONS: INTERVIEWER TURN ON THE TAPE RECORDER AND SAY CLEARLY THE DATE, TIME, LOCATION, AND THE INTERVIEWER’S NAME**

Go through one photo at a time. Ask all these questions for each photo. Whenever a photo is being discussed, explain the photo verbally to the voice recorder

1. **We have [insert number] photos you took which show issues related to your experiences and feelings about water, sanitation and hygiene during or after climate disruptions. Which photo shall we talk about first?**
2. **What is this photo of?**
   - What story are you telling when taking this photo?
   - Why did you take this photo?
   - Why is this important to you?
   - Refer to the topic guide for more questions
3. **What caption would you like to come up with for this photo?**

- *To explain what a caption is, ask the participant to imagine that a person is looking at the photo who doesn’t know them or anything about how they live – their task is to explain to them the experience they were trying to convey in a sentence.*

*When you have discussed each photo:*

4. **Now I’d like you to rank your photos from the least important to most important. Please put the least important issue on the left, and work towards the most important on the right**

- *Take a photo of the ranking, or write the order in your notebook*

5. **Why have you ordered them this way?**

- What makes this the most important issue?
- Run through reasons for the order of all the photos

6. **Those are all of our questions. Do you have anything else you want to say to us?**
7. **Thank you so much for the time you have taken with us. What you have told us will really help us, and others understand the issues faced by people like you. Then we can try and develop approaches to better assist people like you.**

*Leave the printed photos with the participant.*

1. WaterAid (2018) <https://www.wateraid.org/uk/media/teenage-girls-use-photography-to-help-end-the-shame-and-stigma-around-periods-in-nepal>
